# Proteomic profiling of Alzheimer’s disease and Vascular dementia reveals unique underlying signatures

**DOI:** 10.64898/2025.12.08.25341836

**Authors:** Najaf Amin, Pallavi Kaushik, Lazaros Belbasis, Sihao Xiao, M. Austin Argentieri, Shahzad Ahmad, Masud Husain, Rima Kaddurah-Daouk, Cornelia M. van Duijn

## Abstract

**INTRODUCTION:** Alzheimer’s disease (AD) and vascular dementia (VaD) account for most dementia cases. AD biomarkers remain costly and invasive, and no specific biomarkers exist for VaD.

**METHODS:** We analyzed plasma and brain proteomics in the UK Biobank (N=53,000) and ROSMAP (N=512) to identify shared and distinct proteomic signatures of AD and VaD and assess the influence of the APOE ε4 variant.

**RESULTS:** We identified 55 AD-associated and 49 VaD-associated proteins, with 13 shared. AD proteins were enriched in glycosaminoglycan binding and cholesterol metabolism; VaD proteins in virus receptor activity, cytokine activity and metalloproteinases. Both showed IGF pathway dysregulation. APOE ε4 stratification revealed distinct AD proteomic signatures beyond GFAP and NeFL. Mendelian randomization suggested causal links for SNAP25 in AD, EDA2R and TIMP4 in VaD, and PVR in both.

**DISCUSSION:** Findings underscore the importance of APOE genotype and highlight SNAP25, EDA2R, TIMP4, and PVR as potential biomarkers and therapeutic targets.

## 1 Background

Alzheimer’s disease (AD) and vascular dementia (VaD) are the two most common forms of dementia with AD accounting for 60-80% and VaD for 17-20% of all dementia cases, with 10-20% patients exhibiting evidence for both pathologies^1,2^. Recent advances in AD research have led to the National Institute on Aging–Alzheimer’s Association biomarker-based diagnostic framework, which incorporates measures of amyloid-β (Aβ) and tau pathology—assessed through positron emission tomography (PET) or cerebrospinal fluid (CSF)—along with evidence of neurodegeneration in brain regions such as the entorhinal cortex, hippocampus, and temporal lobe on magnetic resonance imaging (MRI)^3^. The past decade has seen major developments in the measurement of biomarkers for AD in blood. Amyloid β (42/40), p-tau-180, p-tau217, glial fibrillary acidic protein (GFAP) and Neurofilament light (NeFL) can be measured in CSF as well as plasma^4^. Plasma p-tau217 has been shown to achieve an accuracy of up to 96% in discriminating AD patients from other forms of dementia^4^. In contrast, GFAP and NeFL serve as general markers of inflammation and neurodegeneration, respectively, and therefore lack specificity for differentiating between dementia subtypes.

VaD is extremely heterogeneous, including small and large vessel disease, ischaemic stroke and intracranial haemorrhage^5,6^. The diagnosis of vascular dementia relies on clinical data and neuroimaging but lacks biomarker-based criteria as developed for AD. Major risk factors of VaD include age, stroke, hypertension, smoking, type 2 diabetes, cardiac disorders, atherosclerosis and metabolic syndrome^3^. Although from an epidemiological perspective one may argue that these risk factors may also contribute to AD patients because of the high prevalence of these risk factors at middle and old age, we hypothesize that the relative contribution may differ between patients who are later diagnosed with AD and those who with vascular dementia. The two dominant risk factors for AD are age and genetic susceptibility. Although about a hundred genetic variants have been identified that increase the risk of AD^7-9^, the epsilon 4 (⍰4) variant of the apolipoprotein E (APOE) gene^2^ is the major genetic driver of the disease risk and age of onset. No major genes have been identified for VaD, mainly due to the lack of well characterized patient-series for genome-wide association studies (GWAS). However, *APOE* ⍰4 shows association with VaD^10^, albeit that the risk of VaD in *APOE* ⍰4 carriers is much lower than that of AD and may reflect the co-occurrence of AD and VaD. Recent proteomic studies have failed to identify unique signatures of VaD as they have been primarily focused on disease prediction^11^. Predictive markers like GFAP, NeFL and GDF15 are not specific and more likely to be a late(r) consequence of the insidious disease^12^. Within AD patients, the early pathogenesis of the disease may depend on the presence of the *APOE* ⍰4 variant. In AD patients, *APOE* ⍰4 not only determines the risk and age at onset of disease but also higher accumulation of tau protein in the brain, atrophy of the medial temporal lobe, and greater deficits in memory, whereas those AD patients who do not carry *APOE* ⍰4 (*APOE* ⍰4^-^) manifest atrophy of the fronto-parietal lobe and greater deficits in non-memory-related cognitive function (e.g., executive function, visuospatial abilities and language)^13^. A recent study performed by the Global Neurodegeneration Proteomics Consortium (GNPC) showed that *APOE* ⍰4^+^ individuals shared a unique proteomic signature irrespective of neurodegenerative diseases^14^.

In the current study, we sought to identify early proteomic signatures of AD and VaD using the data of 53,000 individuals from the UK Biobank profiled for ∼ 3000 proteins with OLINK explore. Within the AD patients, we further stratified the *APOE* ⍰4^+^ and *APOE* ⍰4^-^ cases to identify proteomic signatures of AD that are common and differ across this genetic determinant. We used Mendelian Randomisation to evaluate potentially causal proteins and sought to understand the function of the proteins in the brain using data from the Religious Orders Study/Memory and Aging Project (ROSMAP) study. Study design is illustrated in **Figure 1**.

**Figure 1:**
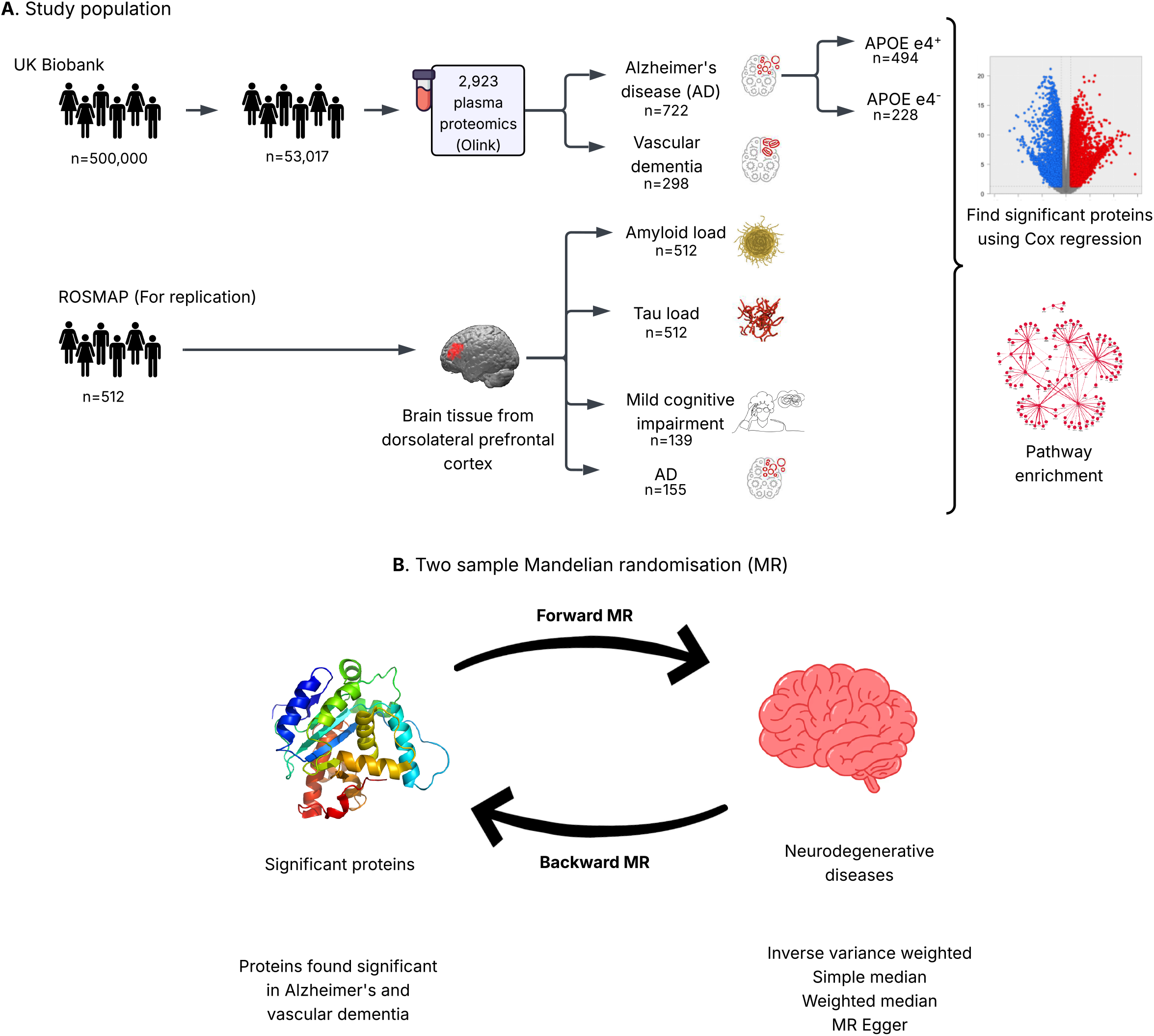
Study design overview. A) Observational discovery analysis using Cox regression in the UK Biobank (plasma) and replication of the significantly associated proteins in the ROSMAP (brain) cohort. B) Causal inference using bi-directional two sample Mendelian Randomisation.

## 2 Methods

### 2.1 Study populations

We performed a prospective, population-based cohort study based on the UK Biobank dataset^15^, which comprised over 500,000 participants aged from 37 to 73 years at recruitment (2006 to 2010). The participants were registered with the UK National Health Service and from 22 assessment centres across England, Wales, and Scotland using standardized procedures for data collection, which included a wide range of questionnaires, anthropological measurement, clinical biomarkers, genotype data, etc. The participants’ hospital inpatient records and death registration were obtained and updated frequently. The updated data until June 2023 were used to define incident diseases in the current study. All participants provided electronically signed informed consent. UK Biobank has approval from the North West Multi-Centre Research Ethics Committee, the Patient Information Advisory Group, and the Community Health Index Advisory Group. Further details on the rationale, study design, survey methods, data collection and ethical approval are available elsewhere^15^. The current study is a part of UK Biobank project 61054.

Brain tissue used in this study was obtained from the autopsy collections of ROSMAP^16^, which is a longitudinal cohort study of aging and dementia in elderly nuns, priests, brothers and lay persons. Brain tissue from the dorsolateral prefrontal cortex used in this study was obtained from the autopsy collections under brain donation programs with standardized protocol^16,17^. The post-mortem neuropathological evaluation, extent of spread of neurofibrillary tangle pathology and neuropathologic diagnoses were made in accordance with established criteria and guidelines^18^. As detailed case metadata with age, sex, post-mortem interval (PMI), APOE genotype, neuropathological criteria and disease status was available in a previous publication^18^. All procedures and research protocols were approved by the corresponding ethical committees of our collaborator’s institutions as well as the Institutional Review Board (IRB) of Columbia University Medical Center (protocol AAAR4962). More details can also be found on the website of Rush Alzheimer’s Disease Center (RADC; https://www.radc.rush.edu/).

### 2.2 Definitions of outcomesd

#### 2.2.1 UK Biobank

We defined incident AD and VaD, and onset dates based on the previous outcome adjudication guidelines in UK Biobank^19^. In brief, the diseases were based on the primary care or the ICD codes from hospital admission electronic health records in the primary or any secondary causes and/or death register. The earliest recorded code date of a disease was used as the date of disease diagnosis. Prevalent cases were defined as the participants with the disease diagnosis date earlier than the first assessment date, reported in the first time self-reported illness. They were excluded from the analysis. The censor date was defined by either the first recorded date of dementia, the death date or the end of the digital recording date, whichever happened first.

#### 2.2.2 ROSMAP

In ROSMAP, we studied two direct neuropathological variables, including overall amyloid levels and the levels of tangle density, which were measured as the mean of the eight brain regions tested. Three derived neuropathological variables were calculated: global neuropsychiatric scores based on the summary of AD pathology derived from counts of three AD pathologies: neuritic plaques, diffuse plaques, and neurofibrillary tangles; AD diagnosed based on the National Institute on Aging (NIA) Reagan score^20^, and neuropsychiatric diagnosis based on Braak and CERAD scores^21^. More details on the study are available on the Rush Alzheimer’s Disease Center website (RADC; https://www.radc.rush.edu/). All subjects gave informed consent.

### 2.3 Covariates used in UK Biobank

The general covariates considered in the analysis included baseline age (field 21022), sex (field 31), BMI (field 21001), fasting time (field 74), assessment center (field 54), spectrometer (field 23650), ethnicity (field 21000), smoking status (field 20116), alcohol intake frequency (field 1558), education (field 6138) and medication use (field 20003) from the verbal interview. Medication status was based on the medication codes collected from the verbal interview, which were coded to Anatomical Therapeutic Chemical (ATC) codes^22^. The medications considered in the covariates were selected based on our previous publication^23^, including five anti-hypertensives (C08, C09, C07, C03 and C02), anti-diabetes (metformin and other anti-diabetes under A10), lipid-lowering drugs (C10), digoxin (C01AA), antithrombotic (B01AC06), proton pump inhibitors (PPI, A02BC), and also 18 drug categories involved in the central nervous system based on 4 digits of the ATC codes.

### 2.4 Genotype measurement in UK Biobank

UK Biobank genotyping was conducted by Affymetrix using a bespoke BiLEVE Axiom array for ∼50K participants and the remaining ∼ 450K on the Affymetrix UK Biobank Axiom array. As the two arrays are broadly comparable with over 95% overlap in assessed gene variants, they were combined. Genetic data was phased prior to imputation with SHAPEIT3 followed by imputation using IMPUTE2. Details on genetic imputation are provided elsewhere^24^. The *APOE* gene (alleles *APOE* ε2, *APOE* ε3, *APOE* ε4) was directly genotyped and defined by 2 single-nucleotide polymorphisms (SNPs), rs429358 and rs7412. Detailed information on the genotyping process and technical methods is available online^25^.

### 2.5 Proteomic profiling

#### 2.5.1 UK Biobank

Proteomic profiling of 54,219 participants from the UK Biobank was performed using the Olink Explore platform that links four Olink panels (Cardiometabolic, Inflammation, Neurology, and Oncology). UK Biobank Olink data are provided as Normalized Protein eXpression (NPX) values on a log2 scale. Details on sample selection, processing, and quality control are provided elsewhere^26^.

#### 2.5.2 ROSMAP

The tandem mass tag (TMT) isobaric labeling mass spectrometry method was used to measure the protein abundance from fresh frozen cortical microdissections of the DLPFC of 618 individuals from ROSMAP. Before TMT labelling, individuals were randomized by covariates (such as age, sex, PMI and diagnosis), into batches (eight individuals per batch). MS/MS (MS2) and SPS-MS3 techniques were used for 45 and five TMT batches via the Orbitrap Fusion mass spectrometer (Thermo Fisher Scientific), respectively. The results were normalized and log2-transformed. Details on sampling, proteomics quantification and quality control are provided elsewhere (https://www.synapse.org/#!Synapse:syn17015098).

### 2.6 Statistical analysis

All analyses were performed in R statistical software (version 4.3.1), and the two-tailed test was considered. Descriptive analysis was performed using the ‘CBCgrps’^27^ library of the R software.

#### 2.6.1 Proteomic signatures of AD and VaD

In the UK Biobank, we used a Cox proportional hazards model to test the relationship between the plasma protein levels at baseline and the risk of incident AD/VaD during the follow-up. A false discovery rate (FDR) of 0.05 was used to identify significance. For the association analysis, we adjusted for a number of covariates including age, sex, BMI, smoking status, alcohol intake frequency, education, ethnicity, physical activity, assessment centre and 27 drugs for common chronic diseases. Additionally, an *APOE* ⍰4 stratified analysis was performed for AD to identify differences between the *APOE* ⍰4^+^ and *APOE* ⍰4^-^ AD patients.

In ROSMAP, linear regression was used to test association of Braak and Cerad scores and logistic regression was used to test the association of MCI and AD with AD/VaD associated proteins in plasma. Analyses were adjusted for post-mortem interval, sex and age.

#### 2.6.2 Protein pathway analysis

Protein pathway analysis was performed using the STRING database. Since we aimed to understand the differences between the two types of dementia including, AD and VaD, and between *APOE* ⍰4^+^ and *APOE* ⍰4^-^ AD cases, we applied an agnostic approach using all genes in the database as the background in pathway analysis.

#### 2.6.3 Mendelian Randomization (MR)

MR analyses were performed using the ‘TwoSampleMR’ library of the R software (version 4.3.1). “Inverse variance weighted” method of MR was considered unless significant directional pleiotropy and/or heterogeneity were observed, in which case results from other methods such as “MR Egger”, “weighted median” and “weighted mode” were used for interpretation. Genetic instruments for AD were extracted from the publicly available genome-wide association study (GWAS) of clinically diagnosed AD by Kunkle et al.^28^. Inferences for VaD were drawn using the GWAS summary statistics from the family history of dementia (FHD) performed in the UKBB and using AD GWAS by Kunkle et al.^28^ as a negative control. The instruments for the proteins were extracted from the summary statistics provided by Sun, B.B et al^26^. For the proteins, we used all independently significantly associated variants (cis & trans, p-value < 10^-11^) as instruments in MR. For other exposures, default settings were used to identify genetic instruments, i.e., p-value < 5^*^10-08 and r2 < 0.001. Steiger test for directionality was used to elucidate causal direction. Leave one out analysis was performed to evaluate the independent contribution of each genetic instrument.

## 3 Results

### 3.1 General characteristics of the study population

Among 53,017 participants whose plasma was profiled for proteomics in the UK Biobank, 722 individuals developed AD and 298 developed VaD over a mean follow-up time of 12.9 years (**Table 1**). Individuals who developed AD and VaD were significantly older and had higher proportions of previous smokers than those who did not develop AD or VaD during follow-up. Individuals who developed VaD had significantly higher BMI, were less active and had a higher proportion of males. Individuals with developed AD or VaD had significantly higher proportions of individuals being treated for diabetes, hypertension, thrombosis, metabolic disorders and dyslipidemia, with the proportion of individuals taking medication being higher in VaD than AD (**Table 1**).

**Table 1:** Descriptive characteristics of the studied population.

| Characteristic | Total<br>(n = 52248) | Controls<br>(n = 51526) | AD<br>(n = 734) | P-value | Total<br>(n = 52244) | Controls (n = 51946) | VAD<br>(n = 298) | P-value |
| --- | --- | --- | --- | --- | --- | --- | --- | --- |
| Recruitment Age, Median (Q1, Q3) | 58 (50, 64) | 58 (50, 63) | 67 (63, 68) | < 0.001 | 58 (50, 64) | 58 (50, 64) | 66 (63, 68) | < 0.001 |
| Sex, n (%) |  |  |  | 0.691 |  |  |  | < 0.001 |
| Female | 28165 (54) | 27764 (54) | 395 (55) |  | 28165 (54) | 28050 (54) | 115 (39) |  |
| Male | 24083 (46) | 23750 (46) | 327 (45) |  | 24079 (46) | 23896 (46) | 183 (61) |  |
| Smoking status, n (%) |  |  |  | < 0.001 |  |  |  | < 0.001 |
| Never | 28277 (54) | 27908 (54) | 362 (50) |  | 28274 (54) | 28150 (54) | 124 (42) |  |
| Previous | 18206 (35) | 17906 (35) | 296 (41) |  | 18205 (35) | 18063 (35) | 142 (48) |  |
| Current | 5512 (11) | 5453 (11) | 59 (8) |  | 5512 (11) | 5482 (11) | 30 (10) |  |
| Alcohol use, n (%) |  |  |  | < 0.001 |  |  |  | < 0.001 |
| Never | 4515 (9) | 4410 (9) | 104 (14) |  | 4515 (9) | 4469 (9) | 46 (15) |  |
| Special occasions only | 6137 (12) | 6029 (12) | 104 (14) |  | 6137 (12) | 6090 (12) | 47 (16) |  |
| One to three times a month | 5686 (11) | 5608 (11) | 78 (11) |  | 5686 (11) | 5656 (11) | 30 (10) |  |
| Once or twice a week | 13516 (26) | 13344 (26) | 170 (24) |  | 13515 (26) | 13451 (26) | 64 (22) |  |
| Three or four times a week | 11749 (23) | 11615 (23) | 133 (18) |  | 11748 (23) | 11699 (23) | 49 (16) |  |
| Daily or almost daily | 10525 (20) | 10390 (20) | 131 (18) |  | 10523 (20) | 10462 (20) | 61 (21) |  |
| BMI, Median (Q1,Q3) | 26.78 (24.18, 29.92) | 26.78 (24.18, 29.92) | 26.88 (24.24, 29.76) | 1 | 26.78 (24.18, 29.92) | 26.78 (24.18, 29.91) | 28.04 (25.18, 31.28) | < 0.001 |
| Physical activity, n (%) |  |  |  | 0.497 |  |  |  | 0.014 |
| low | 8201 (20) | 8104 (20) | 96 (18) |  | 8200 (20) | 8137 (19) | 63 (27) |  |
| moderate | 16943 (40) | 16723 (40) | 216 (40) |  | 16942 (40) | 16859 (40) | 83 (36) |  |
| high | 16834 (40) | 16603 (40) | 227 (42) |  | 16832 (40) | 16745 (40) | 87 (37) |  |
| Ethnic background, n (%) |  |  |  | 0.024 |  |  |  | 0.779 |
| British | 45639 (88) | 44972 (88) | 656 (91) |  | 45635 (88) | 45358 (88) | 277 (93) |  |
| White | 65 (0) | 61 (0) | 3 (0) |  | 65 (0) | 65 (0) | 0 (0) |  |
| Mixed | 1 (0) | 1 (0) | 0 (0) |  | 1 (0) | 1 (0) | 0 (0) |  |
| Asian or Asian British | 5 (0) | 5 (0) | 0 (0) |  | 5 (0) | 5 (0) | 0 (0) |  |
| Black or Black British | 4 (0) | 4 (0) | 0 (0) |  | 4 (0) | 4 (0) | 0 (0) |  |
| Chinese | 146 (0) | 146 (0) | 0 (0) |  | 146 (0) | 146 (0) | 0 (0) |  |
| Other ethnic group | 608 (1) | 604 (1) | 4 (1) |  | 608 (1) | 606 (1) | 2 (1) |  |
| Irish | 1351 (3) | 1329 (3) | 22 (3) |  | 1351 (3) | 1345 (3) | 6 (2) |  |
| Any other white background | 1677 (3) | 1664 (3) | 13 (2) |  | 1677 (3) | 1671 (3) | 6 (2) |  |
| White and Black Caribbean | 75 (0) | 75 (0) | 0 (0) |  | 75 (0) | 75 (0) | 0 (0) |  |
| White and Black African | 62 (0) | 62 (0) | 0 (0) |  | 62 (0) | 62 (0) | 0 (0) |  |
| White and Asian | 91 (0) | 91 (0) | 0 (0) |  | 91 (0) | 91 (0) | 0 (0) |  |
| Any other mixed background | 114 (0) | 111 (0) | 3 (0) |  | 114 (0) | 113 (0) | 1 (0) |  |
| Indian | 576 (1) | 575 (1) | 1 (0) |  | 576 (1) | 575 (1) | 1 (0) |  |
| Pakistani | 171 (0) | 171 (0) | 0 (0) |  | 171 (0) | 171 (0) | 0 (0) |  |
| Bangladeshi | 29 (0) | 29 (0) | 0 (0) |  | 29 (0) | 29 (0) | 0 (0) |  |
| Any other Asian background | 186 (0) | 185 (0) | 1 (0) |  | 186 (0) | 184 (0) | 2 (1) |  |
| Caribbean | 501 (1) | 493 (1) | 8 (1) |  | 501 (1) | 500 (1) | 1 (0) |  |
| African | 680 (1) | 673 (1) | 7 (1) |  | 680 (1) | 679 (1) | 1 (0) |  |
| Any other Black background | 13 (0) | 13 (0) | 0 (0) |  | 13 (0) | 13 (0) | 0 (0) |  |
| APOE, n (%) |  |  |  | < 0.001 |  |  |  | < 0.001 |
| e22 | 431 (1) | 427 (1) | 4 (1) |  | 431 (1) | 428 (1) | 3 (1) |  |
| e23 | 6334 (12) | 6304 (12) | 30 (4) |  | 6334 (12) | 6323 (12) | 11 (4) |  |
| e24 | 1382 (3) | 1369 (3) | 12 (2) |  | 1382 (3) | 1372 (3) | 10 (3) |  |
| e33 | 30371 (58) | 30171 (59) | 194 (27) |  | 30369 (58) | 30248 (58) | 121 (41) |  |
| e34 | 12248 (23) | 11931 (23) | 312 (43) |  | 12246 (23) | 12134 (23) | 112 (38) |  |
| e44 | 1480 (3) | 1310 (3) | 170 (24) |  | 1480 (3) | 1439 (3) | 41 (14) |  |
| mutation | 2 (0) | 2 (0) | 0 (0) |  | 2 (0) | 2 (0) | 0 (0) |  |
| Survival time (days), Median (Q1, Q3) | 4904 (4643, 5147) | 4910 (4654, 5150) | 3215.5 (2036.5, 4309.5) | < 0.001 | 4906 (4648, 5149) | 4909 (4652, 5150) | 3497.5 (2220.5, 4243.75) | < 0.001 |
| Antidiabetics, n (%) |  |  |  | < 0.001 |  |  |  | < 0.001 |
| No | 50085 (96) | 49422 (96) | 651 (90) |  | 50081 (96) | 49824 (96) | 257 (86) |  |
| Yes | 2163 (4) | 2092 (4) | 71 (10) |  | 2163 (4) | 2122 (4) | 41 (14) |  |
| Antihypertensives, n (%) |  |  |  | 0.1 |  |  |  | 0.002 |
| No | 51385 (98) | 50670 (98) | 704 (98) |  | 51381 (98) | 51096 (98) | 285 (96) |  |
| Yes | 863 (2) | 844 (2) | 18 (2) |  | 863 (2) | 850 (2) | 13 (4) |  |
| Diuretics, n (%) |  |  |  | < 0.001 |  |  |  | < 0.001 |
| No | 47702 (91) | 47072 (91) | 619 (86) |  | 47699 (91) | 47463 (91) | 236 (79) |  |
| Yes | 4546 (9) | 4442 (9) | 103 (14) |  | 4545 (9) | 4483 (9) | 62 (21) |  |
| Beta blockers, n (%) |  |  |  | < 0.001 |  |  |  | < 0.001 |
| No | 48257 (92) | 47622 (92) | 626 (87) |  | 48253 (92) | 48020 (92) | 233 (78) |  |
| Yes | 3991 (8) | 3892 (8) | 96 (13) |  | 3991 (8) | 3926 (8) | 65 (22) |  |
| Calcium channel blockers, n (%) |  |  |  | < 0.001 |  |  |  | < 0.001 |
| No | 48096 (92) | 47477 (92) | 607 (84) |  | 48094 (92) | 47861 (92) | 233 (78) |  |
| Yes | 4152 (8) | 4037 (8) | 115 (16) |  | 4150 (8) | 4085 (8) | 65 (22) |  |
| RAS modifying agents, n (%) |  |  |  | < 0.001 |  |  |  | < 0.001 |
| No | 44290 (85) | 43756 (85) | 525 (73) |  | 44286 (85) | 44093 (85) | 193 (65) |  |
| Yes | 7958 (15) | 7758 (15) | 197 (27) |  | 7958 (15) | 7853 (15) | 105 (35) |  |
| Lipid modifying agents, n (%) |  |  |  | < 0.001 |  |  |  | < 0.001 |
| No | 41197 (79) | 40771 (79) | 419 (58) |  | 41195 (79) | 41048 (79) | 147 (49) |  |
| Yes | 11051 (21) | 10743 (21) | 303 (42) |  | 11049 (21) | 10898 (21) | 151 (51) |  |
| Opioids, n (%) |  |  |  | < 0.001 |  |  |  | < 0.001 |
| No | 49285 (94) | 48622 (94) | 652 (90) |  | 49281 (94) | 49018 (94) | 263 (88) |  |
| Yes | 2963 (6) | 2892 (6) | 70 (10) |  | 2963 (6) | 2928 (6) | 35 (12) |  |
| Analgesics/antipyretics, n (%) |  |  |  | < 0.001 |  |  |  | < 0.001 |
| No | 35999 (69) | 35576 (69) | 418 (58) |  | 35998 (69) | 35866 (69) | 132 (44) |  |
| Yes | 16249 (31) | 15938 (31) | 304 (42) |  | 16246 (31) | 16080 (31) | 166 (56) |  |
| Antimigraine, n (%) |  |  |  | 0.014 |  |  |  | 1 |
| No | 51623 (99) | 50890 (99) | 721 (100) |  | 51619 (99) | 51324 (99) | 295 (99) |  |
| Yes | 625 (1) | 624 (1) | 1 (0) |  | 625 (1) | 622 (1) | 3 (1) |  |
| Antiepileptics, n (%) |  |  |  | < 0.001 |  |  |  | < 0.001 |
| No | 51182 (98) | 50482 (98) | 690 (96) |  | 51178 (98) | 50897 (98) | 281 (94) |  |
| Yes | 1066 (2) | 1032 (2) | 32 (4) |  | 1066 (2) | 1049 (2) | 17 (6) |  |
| Anticholinergic agents, n (%) |  |  |  | 0.403 |  |  |  | 0.191 |
| No | 52211 (100) | 51478 (100) | 721 (100) |  | 52207 (100) | 51910 (100) | 297 (100) |  |
| Yes | 37 (0) | 36 (0) | 1 (0) |  | 37 (0) | 36 (0) | 1 (0) |  |
| Dopaminergic agents, n (%) |  |  |  | 0.111 |  |  |  | 0.631 |
| No | 52065 (100) | 51336 (100) | 717 (99) |  | 52061 (100) | 51763 (100) | 298 (100) |  |
| Yes | 183 (0) | 178 (0) | 5 (1) |  | 183 (0) | 183 (0) | 0 (0) |  |
| Antipsychotics, n (%) |  |  |  | 0.08 |  |  |  | < 0.001 |
| No | 51746 (99) | 51024 (99) | 710 (98) |  | 51743 (99) | 51455 (99) | 288 (97) |  |
| Yes | 502 (1) | 490 (1) | 12 (2) |  | 501 (1) | 491 (1) | 10 (3) |  |
| Anxiolytics, n (%) |  |  |  | < 0.001 |  |  |  | < 0.001 |
| No | 51977 (99) | 51256 (99) | 709 (98) |  | 51973 (99) | 51683 (99) | 290 (97) |  |
| Yes | 271 (1) | 258 (1) | 13 (2) |  | 271 (1) | 263 (1) | 8 (3) |  |
| Hypnotics/sedatives, n (%) |  |  |  | 0.647 |  |  |  | 0.083 |
| No | 51844 (99) | 51115 (99) | 718 (99) |  | 51840 (99) | 51547 (99) | 293 (98) |  |
| Yes | 404 (1) | 399 (1) | 4 (1) |  | 404 (1) | 399 (1) | 5 (2) |  |
| Antidepressants, n (%) |  |  |  | < 0.001 |  |  |  | < 0.001 |
| No | 48096 (92) | 47463 (92) | 625 (87) |  | 48093 (92) | 47846 (92) | 247 (83) |  |
| Yes | 4152 (8) | 4051 (8) | 97 (13) |  | 4151 (8) | 4100 (8) | 51 (17) |  |
| Psychostimulants, n (%) |  |  |  | 1 |  |  |  | 1 |
| No | 52229 (100) | 51495 (100) | 722 (100) |  | 52225 (100) | 51927 (100) | 298 (100) |  |
| Yes | 19 (0) | 19 (0) | 0 (0) |  | 19 (0) | 19 (0) | 0 (0) |  |
| Psycholeptics & psychoanaleptics, n (%) |  |  |  | 1 |  |  |  | 1 |
| No | 52227 (100) | 51493 (100) | 722 (100) |  | 52223 (100) | 51925 (100) | 298 (100) |  |
| Yes | 21 (0) | 21 (0) | 0 (0) |  | 21 (0) | 21 (0) | 0 (0) |  |
| Antidementia drugs, n (%) |  |  |  | < 0.001 |  |  |  | 0.082 |
| No | 51962 (99) | 51257 (100) | 698 (97) |  | 51958 (99) | 51664 (99) | 294 (99) |  |
| Yes | 286 (1) | 257 (0) | 24 (3) |  | 286 (1) | 282 (1) | 4 (1) |  |
| Parasympathomimetics, n (%) |  |  |  | 1 |  |  |  | 1 |
| No | 52 223 (100) | 51 489 (100) | 722 (100) |  | 52 219 (100) | 51 921 (100) | 298 (100) |  |
| Yes | 25 (0) | 25 (0) | 0 (0) |  | 25 (0) | 25 (0) | 0 (0) |  |
| Antiaddiction drugs, n (%) |  |  |  | 0.088 |  |  |  | 0.353 |
| No | 52 172 (100) | 51 441 (100) | 719 (100) |  | 52 168 (100) | 51 871 (100) | 297 (100) |  |
| Yes | 76 (0) | 73 (0) | 3 (0) |  | 76 (0) | 75 (0) | 1 (0) |  |
| Antivertigo, n (%) |  |  |  | 0.037 |  |  |  | 0.366 |
| No | 52 024 (100) | 51 297 (100) | 715 (99) |  | 52 020 (100) | 51 724 (100) | 296 (99) |  |
| Yes | 224 (0) | 217 (0) | 7 (1) |  | 224 (0) | 222 (0) | 2 (1) |  |
| Anesthetics, n (%) |  |  |  | 1 |  |  |  | 1 |
| No | 52 223 (100) | 51 489 (100) | 722 (100) |  | 52 219 (100) | 51 921 (100) | 298 (100) |  |
| Yes | 25 (0) | 25 (0) | 0 (0) |  | 25 (0) | 25 (0) | 0 (0) |  |
| Proton pump inhibitors, n (%) |  |  |  | < 0.001 |  |  |  | < 0.001 |
| No | 46 579 (89) | 45 991 (89) | 579 (80) |  | 46 575 (89) | 46 355 (89) | 220 (74) |  |
| Yes | 5 669 (11) | 5 523 (11) | 143 (20) |  | 5 669 (11) | 5 591 (11) | 78 (26) |  |
| Digitalis glycosides, n (%) |  |  |  | 0.136 |  |  |  | 0.009 |
| No | 52 024 (100) | 51 296 (100) | 716 (99) |  | 52 021 (100) | 51 728 (100) | 293 (98) |  |
| Yes | 224 (0) | 218 (0) | 6 (1) |  | 223 (0) | 218 (0) | 5 (2) |  |
| Platelet aggregation inhibitors, n (%) |  |  |  | < 0.001 |  |  |  | < 0.001 |
| No | 44 290 (85) | 43 755 (85) | 528 (73) |  | 44 289 (85) | 44 103 (85) | 186 (62) |  |
| Yes | 7 958 (15) | 7 759 (15) | 194 (27) |  | 7 955 (15) | 7 843 (15) | 112 (38) |  |

### 3.2 Proteomic signatures of AD in plasma

AD was significantly associated (false discovery rate (FDR) < 0.05) with 55 proteins, including GFAP (Beta=1.02, FDR=9.0^*^10^-77^), NEFL (Beta=0.69, FDR=5.7^*^10^-20^) and APOE (Beta=-1.2, FDR=4.2^*^10^-35^) showing the most statistically significant associations (**Figure 2, Supplementary Table 1**). Other plasma proteins strongly associated with AD include MENT, SNAP25, SYT1, VGF, NPTXR, CEND1. The correlation structure of the 55 proteins shows no strong correlations among proteins in either those who developed AD during follow-up or those who did not (**Supplementary Figures 1a and 1b**). Pathway analysis suggests that these proteins are involved in glycosaminoglycan binding (GO), cholesterol metabolism (KEGG) and regulation of Insulin-like growth factor (IGF) transport and uptake (Reactome) among others (**Supplementary Table 2**).

**Figure 2:**
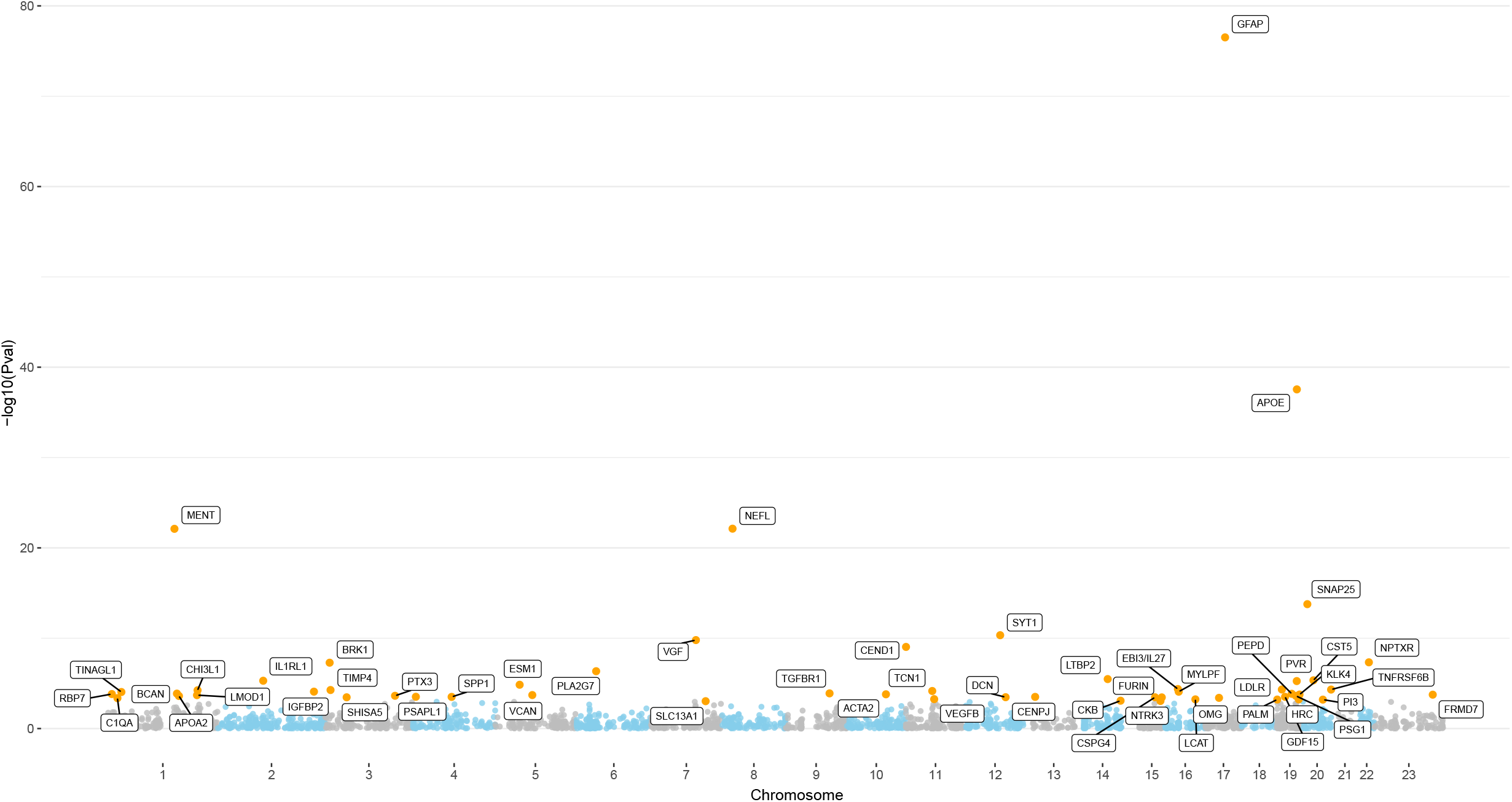
Proteome-wide association plot of AD. Each dot represents a protein where FDR significant proteins are highlighted in yellow. X-axis shows the chromosomes which the genes coding the proteins are mapped to. Y-axis depicts the strength of the association.

### 3.3 Proteomic signatures of AD show no overlap between *APOE* ⍰4+ patients and APOE ⍰4-patients beyond GFAP & NeFL

When stratified by *APOE* ⍰4 status, *APOE* ⍰4^+^ and *APOE* ⍰4^-^, the two series of AD cases showed different proteomic signatures beyond GFAP and NEFL despite a significant correlation (r = 0.295, p-value = 1.2^*^10^-59^) (**Supplementary Table 3, Figure 3**). In *APOE* ⍰4^+^ individuals, AD was associated with 15 proteins: GFAP, NEFL, VGF, SYT1, CEND1, MENT, NPTXR, LDLR, RBP7, IL1RL1, IL27, TNFRSF6B, SLC13A1, PSG1 and STAB2. In *APOE* ⍰4^-^ individuals AD was associated with 24 proteins: GFAP, NEFL, FRMD7, CST5, VEGFB, TGFBR1, ADGRD1, SMOC2, GFRA2, CELSR2, CTHRC1, SHISA5, YAP1, GUCY2C, BMPER, CLSTN2, LTBP2, IL24, DTX3, FLT1, KLK13, LMOD1, NPTX2 and FBLN2. Proteins including ADGRD1, IL24, CTHRC1, CELSR2 are particularly interesting as those not only show no association with AD in *APOE* ⍰4^+^ but also show opposite effects (**Figure 3**). Of note is that also the plasma levels of the MENT protein are in the opposite direction when comparing *APOE* ⍰4^+^ and *APOE* ⍰4^-^ carriers, being significantly associated with future AD in *APOE* ⍰4^+^ carriers.

**Figure 3:**
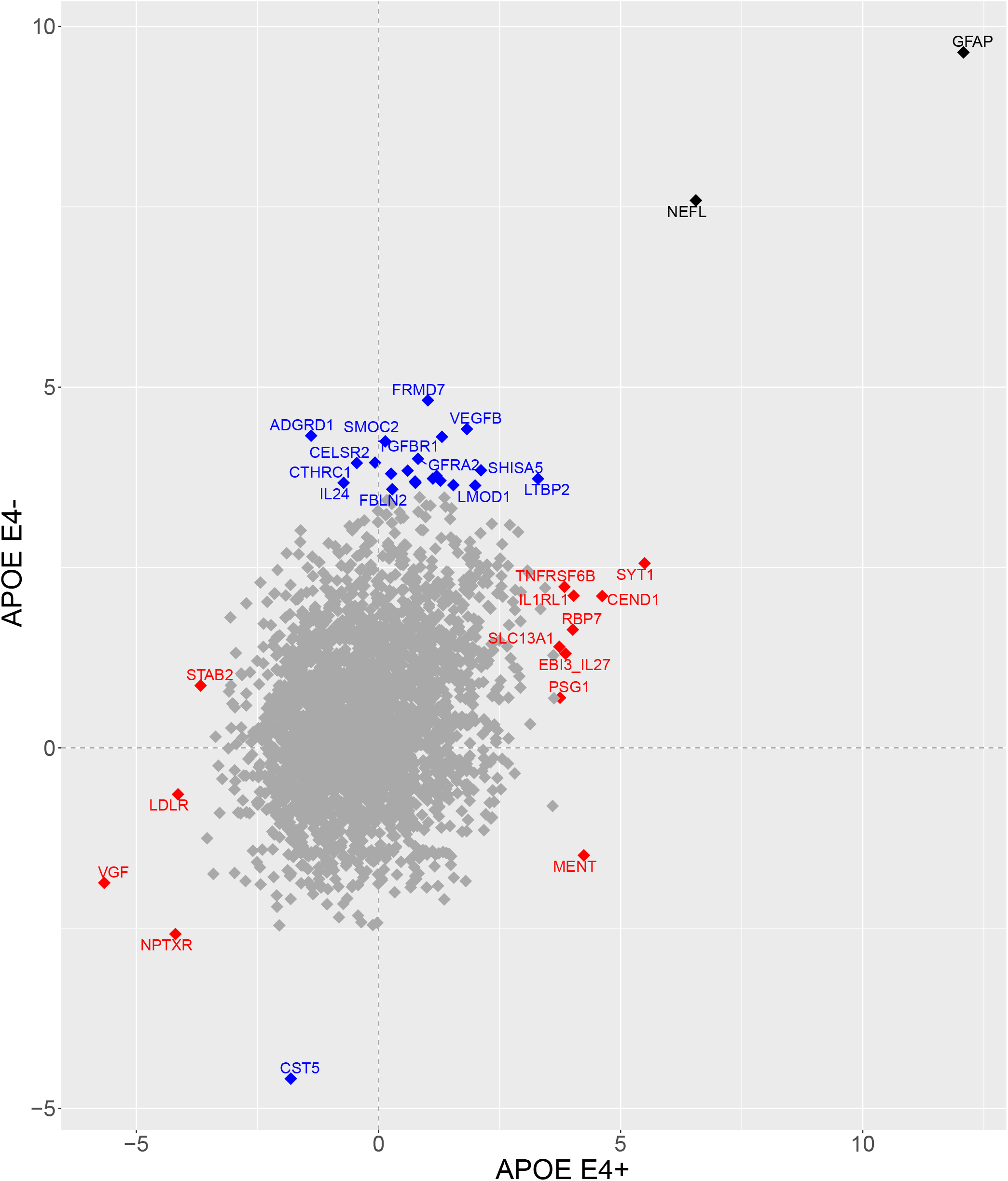
Scatter plot of proteomic signatures of AD (*APOE* ⍰4^+^) vs AD (*APOE* ⍰4^-^). Axes depict the Z-scores from proteome-wide association analyses in the two strata. Each diamond represents a protein, where FDR significant ones are highlighted in blue, red and black. Blue are the proteins uniquely associated with AD in the *APOE* ⍰4^-^ group and red are the proteins uniquely associated with AD in the *APOE* ⍰4^+^ group. Black diamonds are the proteins associated with AD in both groups.

Together, the stratified analyses leaves 21 proteins of the 55 proteins identified in the overall analysis including VGF, SYT1, CEND1, MENT, NPTXR, LDLR, IL1RL1, RBP7, EBI3-IL27, TNFRSF6B, SLC13A1 and PSG1 which were significantly associated with AD only in *APOE* ⍰4^+^, LTBP2, SHISA5, CST5, VEGFB, LMOD, TGFBR1 and FRMD7 which were significantly associated with AD in *APOE* ⍰4^-^ carriers, and GFAP and NEFL, which were associated with AD in both *APOE* ⍰4^+^ and *APOE* ⍰4^-^ carriers (**Supplementary Figure 2**). There were 16 proteins we additionally identified in the stratified analysis, mostly (15/16) in the *APOE* ⍰4^-^ group. These include NPTX2, ADGRD1, DTX3, BMPER, ALSTN2, GFRA2, FLT1, KLK13, IL24, YAP1, CTHRC1, FNLN2, GUCY2C, SMOC2 and CELSR2 and one protein STAB2 in the *APOE* ⍰4^+^ carriers.

Proteins associated with AD in *APOE* ⍰4^+^ carriers localize to the endocytic vesicle membrane, and are involved in lipoprotein binding and chylomicron clearance (**Supplementary Table 4**). In contrast, most of the proteins associated with AD in *APOE* ⍰4^-^ individuals localize to the extracellular region and do not cluster in any specific pathway (**Supplementary Table 5**).

### 3.4 The CELSR2 effect in those without APOE lll4 variant is confirmed in a APOE stratified GWAS analysis

CELSR2 belongs to the flamingo family of cadherins and is encoded by the gene *CELSR2*, which hosts the genetic variant (rs12740374_T, 3’UTR) that determines the plasma levels of progranulin (GRN, effect=-0.664, p-value=0)^26^ (**Supplementary Figure 3**). CELSR2 localizes to the same chromosomal region as SORT1 (**Supplementary Figure 3**), which encodes sortilin 1 protein that is primarily responsible for the degradation of GRN^29^. Low progranulin levels in the brain are known to be causal for several types of dementia, including frontotemporal lobar degeneration (FTLD) and amyotrophic lateral sclerosis (ALS) but in AD patients, higher CSF levels of progranulin have been associated with disease progression^30^. However, previous MR studies suggest a causal association between plasma GRN and AD^30^. In both the AD overall analysis and in the *APOE* stratified analysis, plasma levels of GRN and SORT1 proteins showed no association with the risk of future AD. In this context another interesting plasma protein is CST5, which is one of the most significant proteins in *APOE* ⍰4^-^ (Beta= - 0.61, FDR= 3.3^*^10^-03^) but does not show any association with AD in APOE ⍰4^+^ (Beta= -0.16, FDR=0.48). CST5 encodes cystatin D, a protein that interacts with GRN by inhibiting cathepsin S and through GRN affects the expression of several mitochondrial transporters (**Supplementary Figure 4**). Together, our findings in blood suggest that the relevance of progranulin to AD may be specific to *APOE* ⍰4^-^ AD patients. This finding is consistent with that of *APOE* ⍰4 stratified GWAS^31^ where the GRN locus shows genome-wide significant association with AD only in the *APOE* ⍰4^-^ group.

### 3.5 Proteomic signatures of VaD suggest dysregulation of the virus receptor pathway

VaD showed a significant association (FDR < 0.05) with 49 proteins in plasma, including GFAP, NEFL, CHI3L1, PVR, GDF15, SPON2, SMOC2, C7, CKB, ASAH2 and BSG among others (**Figure 4, Supplementary Table 6**). Of note is that the APOE protein in plasma emerges as significantly associated to the future risk of VaD (Beta= -0.40, FDR=0.05), however, the significance and strength is not comparable to that seen in AD (Beta= -1.2, FDR=4.2^*^10^-35^). Pathway enrichment analysis suggests that the proteins with the future risk of VaD are enriched in the virus receptor, cytokine activity, matrix metalloproteinases and regulation of Insulin-like Growth Factor (IGF) transport and uptake pathways (**Supplementary Table 7**).

**Figure 4:**
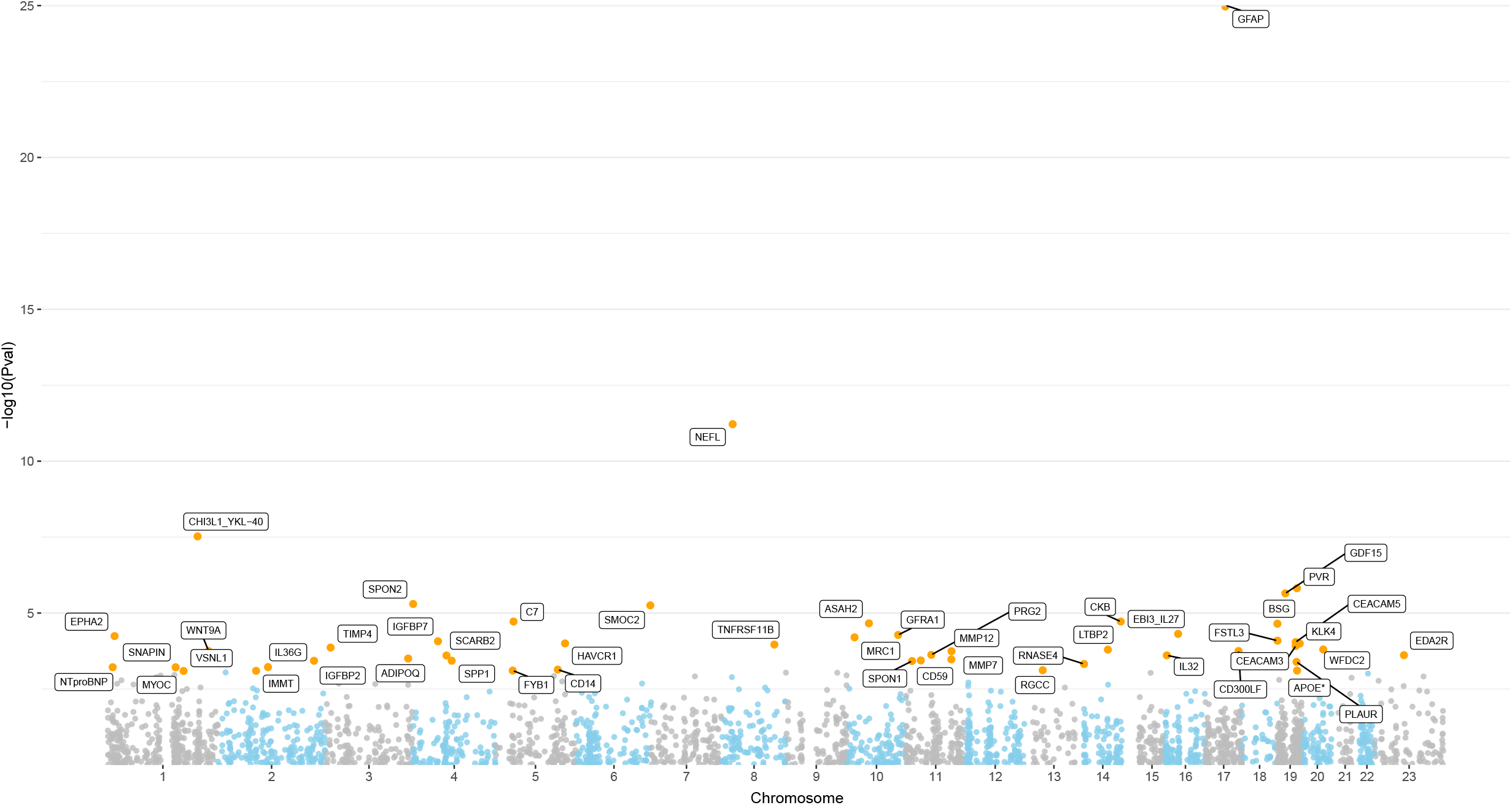
Proteome-wide association plot of VaD. Each dot represents a protein where FDR significant are highlighted in yellow. X-axis shows the chromosomes which the genes coding the proteins are mapped to. Y-axis depicts the strength of the association.

When comparing the proteomic signatures of VaD with that of the AD, a significant correlation was observed (r=0.64, p-value = 7.15^*^10^-314^) (**Supplementary Figure 5**). However, among significant ones 13/49 proteins including GFAP, NEFL, IL27, IGFBP2, LTBP2, APOE, KLK4, PVR, CKB, SPP1, CHI3L1, GDF15 and TIMP4 overlap with that of AD. 11/13 overlapping proteins are localized to the extracellular region, of which 3 cluster in elastic fibre formation and matrix metalloproteinases pathway (**Supplementary Table 8**).

The virus receptor pathway appears to be most prominent in those who develop VaD in the future. This cluster includes 6 proteins: PVR, HAVCR1, SCARB2, BSG, EPHA2 and MRC1. Of these only PVR showed a significant association with AD. HAVCR1 has been associated with family history of dementia in a GWAS^32^ but not with AD. Correlation structure of the 49 proteins associated with VaD shows a strongly correlated cluster of 12 proteins including SPON2, SMOC2, GDF15, PLAUR, WFDC2, EDA2R, EPHA2, BSG, GFRA1, SCARB2, FSTL3 and CD59 particularly among cases (**Supplementary Figure 6**). When comparing the significantly associated proteins in AD overall, AD-*APOE* ⍰4^+^, AD-*APOE* ⍰4^-^ and VaD, VaD clustered more closely with AD-*APOE* ⍰4^-^ (**Supplementary Figure 7**).

### 3.6 Virus receptors and the risks of Varicella Zoster (VZV) and Herpes Simplex (HSV1) viral infections

Varicella Zoster (VZV) and herpes simplex virus (HSV1) infections have been consistently associated with increased risk of dementia^33^. The association of virus receptors particularly with VaD prompted us to test the association of these proteins and the seropositivity of VZV and HSV1 in a subset of individuals from the UK Biobank who were characterized for both seropositivity of various viral infections and proteomics (N=967). Proteins PVR and EPHA2 were associated with seropositivity of VZV, while HAVCR1 was associated with seropositivity of HSV1 (Supplementary Table 9). All six proteins were significantly associated with the two established markers of neurodegeneration, i.e., GFAP and NEFL (Supplementary Table 10). 3.7 Replication of proteomic signatures of AD in and VaD in the brain (ROSMAP)

To evaluate the relevance of plasma proteins associated with AD and VaD in neurodegeneration in the brain, we tested their association with neuritic plaques based on the Consortium to Establish a Registry for Alzheimer’s Disease (CERAD) score, tau pathology as measured with the Braak stage, mild cognitive impairment (MCI) and AD in the ROSMAP cohort (Supplementary Table 11). Of the 55 proteins associated with AD in plasma, 31 were assessed in the ROSMAP dataset. Several proteins, including GFAP, NEFL, VGF, APOE, SNAP25, PVR, VCAN, LMOD1, DCN, PEPD, PALM, CHI3L1, TINAGL1, ACTA2 and SYT1, were associated with at least one trait tested (Supplementary Table 12). SNAP25 was particularly interesting as it was significantly associated with MCI (beta = -3.75, p-value = 3.9^*^10^-04^), tau pathology (beta= -1.71, p-value = 1.4^*^10^-04^), neuritic plaques (beta = -1.86, p-value = 9.7^*^10^-06^) and AD (beta = -3.49, p-value = 2.9^*^10^-05^). In a previous study, cerebrospinal fluid levels of SNAP25 were found to be associated with MCI and AD^34^ and another study by Wingo et al^35^., PVR was identified as the top protein associated with AD independent of APOE E4 in brain but was not followed up as the protein was absent in the replication dataset.

Of the 36 unique proteins associated with VaD in plasma, 15 were assessed in the ROSMAP dataset, of which IGFBP7, RGCC, IMMT, BSG, SPON1, RNASE4 and SNAPIN showed association with at least one of the tested traits (Supplementary Table 13). Among these SPON1 is particularly interesting as it showed a very strong significant association with MCI (beta= 2.66, p-value = 4.0^*^10^-03^), tau pathology (beta = 1.99, p-value= 1.5^*^10^-06^), neuritic plaques (beta=2.42, p-value = 3.1^*^10^-10^) but not with AD, suggesting that it may be specific to VaD.

### 3.8 Mendelian Randomization analysis identifies specific and common proteins influencing the risk of AD & VaD

We used publicly available summary statistics from Sun et al. 2023^26^ for proteins. We selected all independent genome-wide significant SNPs (*cis & trans*) as instrumental variables (https://metabolomics.helmholtz-munich.de/ukbbpgwas/pgwas.table.php). We used summary statistics from Kunkle et al.^28^ to run MR analysis for AD since it is the largest GWAS published that includes only diagnosed patients of AD. Since there are no known genetic associations for VaD, we used summary statistics from GWAS performed on the family history of dementia (FHD) in the UKBB to draw inferences for VaD. Among the proteins significantly associated with AD in the Cox regression analysis, APOE, SNAP25, PVR and PALM showed significant association in MR (**Supplementary Tables 14 & 15**). The MR association with APOE, SNAP25 and PALM was driven by the *APOE* ⍰4 variant rs429358-C (**Supplementary Table 16, Supplementary Figure 8**). Of these three, SNAP25 showed the strongest association with early AD pathology in the brain and the protein is directly associated to APOE and expressed in particular in the cingulate cortex while the association between PALM and APOE is indirect through SNAP25 (**Supplementary Figure 9**), we therefore chose SNAP25 as the primary causal candidate for AD. Since the *APOE* ⍰4 variant rs429358-C is a pleiotropic variant, violating the assumption of MR, we further used MR-Egger to draw causal inferences in the presence of pleiotropy. MR-Egger showed a significant causal association of SNAP25 with AD (Beta = 2.12, P-value = 1.6^*^10^-04^; **Supplementary Table 14**) after correcting for directional pleiotropy. The causal effect was stronger than the Egger intercept, suggesting that the Egger estimate is unbiased (**Table 2**). Further, the results of MR methods, e.g., weighted median and mode, were also significant and consistent with that of the inverse variance weighted method (**Supplementary Table 14**). Genetically, PVR is an independent locus in the neighbourhood of APOE with relatively weak LD to APOE (**Supplementary Figure 10**). In a previous study, PVR was also found to be causally associated with AD in the brain independent of APOE genotype^35^.

**Table 2:** Results of MR analysis for the selected proteins.

|  |  |  | Inverse Variance |  |  | Pleiotropy |  |  | Causal Direction |  |  |  |
| --- | --- | --- | --- | --- | --- | --- | --- | --- | --- | --- | --- | --- |
| Exposure | outcome | Nsnps | Beta | SE | Pval | Intercept | SE | pval | Exposure R2 | Outcome R2 | Direction | pval |
| **Forward MR** |  |  |  |  |  |  |  |  |  |  |  |  |
| SNAP25 | AD | 6 | 1.767 | 0.241 | 0.00E+00 | -0.1398 | 0.0362 | 1.81E-02 | 0.053 | 0.014 | TRUE | 0 |
| SNAP25 | FHD | 11 | 0.745 | 0.095 | 0.00E+00 | -0.0599 | 0.0094 | 1.29E-04 | 0.056 | 3.18E-03 | TRUE | 0 |
| PVR | AD | 2 | 0.140 | 0.022 | 0.00E+00 | - | - | - | 0.021 | 6.72E-04 | TRUE | 0 |
| PVR | FHD | 4 | 0.061 | 0.010 | 0.00E+00 | -0.0021 | 0.0082 | 8.23E-01 | 0.023 | 1.36E-04 | TRUE | 0 |
| PVR | VZV | 4 | 0.066 | 0.081 | 4.12E-01 | 0.0078 | 0.0677 | 9.18E-01 | 0.023 | 1.12E-04 | TRUE | 0 |
| EDA2R | AD | 10 | 0.170 | 0.128 | 1.82E-01 | 0.0023 | 0.0146 | 8.77E-01 | 0.013 | 9.22E-05 | TRUE | 0 |
| EDA2R | FHD | 10 | 0.164 | 0.055 | 3.13E-03 | 0.0003 | 0.0068 | 9.68E-01 | 0.013 | 5.95E-05 | TRUE | 0 |
| EDA2R | VZV | 10 | 1.061 | 0.457 | 2.02E-02 | -0.0069 | 0.0523 | 8.99E-01 | 0.013 | 7.61E-04 | TRUE | 0 |
| TIMP4 | AD | 6 | -0.047 | 0.045 | 3.01E-01 | -0.0007 | 0.0122 | 9.54E-01 | 0.027 | 1.17E-04 | TRUE | 0 |
| TIMP4 | FHD | 10 | 0.065 | 0.021 | 2.53E-03 | -0.0042 | 0.0038 | 2.99E-01 | 0.032 | 8.20E-05 | TRUE | 0 |
| TIMP4 | VZV | 10 | 0.379 | 0.181 | 3.64E-02 | -0.0284 | 0.0324 | 4.06E-01 | 0.032 | 1.79E-03 | TRUE | 0 |
| **Reverse MR** |  |  |  |  |  |  |  |  |  |  |  |  |
| AD | SNAP25 | 23 | 0.958 | 0.098 | 2.19E-22 | -0.0520 | 0.0065 | 8.98E-08 | 1.92E-03 | 0.015 | FALSE | 4.29E-39 |
| AD | PVR | 23 | 0.093 | 0.017 | 1.13E-07 | -0.0041 | 0.0019 | 4.71E-02 | 1.92E-03 | 9.44E-07 | TRUE | 2.43E-14 |
| FHAD | PVR | 14 | 0.118 | 0.028 | 3.40E-05 | -0.0023 | 0.0037 | 5.46E-01 | 5.74E-05 | 7.60E-07 | TRUE | 0.16 |
| FHAD | EDA2R | 14 | -0.052 | 0.016 | 1.64E-03 | 0.0023 | 0.0024 | 3.39E-01 | 5.74E-05 | 7.70E-08 | TRUE | 0.13 |
| FHAD | TIMP4 | 14 | 0.011 | 0.026 | 6.77E-01 | 0.0033 | 0.0037 | 3.90E-01 | 5.74E-05 | 7.01E-08 | TRUE | 0.13 |

Among the proteins significantly and specifically associated with VaD in observational analysis, EDA2R and TIMP4 showed significant MR results with FHD but not with AD suggesting that these two proteins may be specific to VaD. Interestingly both proteins were also causally associated with VZV seropositivity (**Supplementary Tables 14 & 15**).

For our prospective causal candidates, i.e., SNAP25, PVR, EDA2R and TIMP4 we further performed a reverse MR, i.e., with AD/FHD as the exposure and protein plasma levels as the outcome, to confirm the causal direction. SNAP25 showed significant MR, but the Steiger test for directionality suggests that changes in SNAP25 levels are upstream of the disease process (R^2^_AD_ = 0.0019 vs R^2^_SNAP25_ = 0.015; p-value = 4.29^*^10^-39^) (**Table 2**). The leave-one-out analysis suggests that the APOE variant rs429358-C is the primary driver of MR association (**Supplementary Figure 11**). Both AD and FHD showed a significant causal association with plasma levels of PVR, but the significance was driven only by the APOE variant (rs429358-C) for AD and a rare variant rs12972156 in the APOE region for FHD (**Supplementary Figure 12**). For VaD-specific candidate proteins, FHD was not causally associated with TIMP4 plasma levels, and EDA2R showed a significant association but inconsistent direction compared to association analysis (**Table 2**). These results reiterate that the plasma levels of SNAP25 may be causal for AD, while plasma levels of EDA2R and TIMP4 may be causally associated with VaD and plasma levels of PVR may be causal for dementia.

## 4 Discussion

In this study, we identified 55 proteins associated with AD and 49 with VaD, with 13 overlapping proteins primarily localized to the extracellular region. AD-associated proteins were uniquely enriched in glycosaminoglycan synthesis and cholesterol metabolism, whereas VaD-associated proteins were enriched in virus receptor and cytokine activity. Both diseases showed dysregulation of the IGF transport and uptake pathway. Stratifying by *APOE* ⍰4 status revealed distinct AD proteomic signatures: *APOE* ⍰4 carriers showed differential levels of proteins on endocytic vesicle membranes involved in lipoprotein binding and chylomicron clearance, while non-carriers showed changes in extracellular proteins without specific pathway enrichment. Mendelian randomization suggests that high plasma SNAP25 levels driven by the *APOE* ⍰4 variant may contribute to AD pathogenesis, while elevated EDA2R and TIMP4 may be causally linked to VaD. High PVR levels may contribute to both AD and VaD. SNAP25 was significantly associated with MCI, AD, and AD-related brain pathology, and PVR with AD. Among VaD-associated proteins replicated in ROSMAP, SPON1 was notable for its strong associations with MCI, tau pathology, and neuritic plaques, but not AD, suggesting specificity for VaD.

Synaptosomal-Associated Protein 25 (SNAP25) is the most interesting finding for AD. The protein is a component of the SNARE complex involved in the exocytotic release of neurotransmitters during synaptic transmission^36^. SNAP25 is highly expressed in the hippocampal neurons^37^. Genetic variants in *SNAP25* gene have previously been associated with cognitive ability in healthy individuals^38^, AD and MCI^39^. Further, high CSF levels of SNAP25 have been associated with MCI, AD and Cruetzfeldt Jacob Disease but not with ALS, FTLD or with Parkinson’s disease (PD)^34^. In our study, we find that SNAP25 levels are significantly increased in the plasma of future AD patients and significantly decreased in the brains of those who have MCI and AD. Plasma levels of SNAP25 are mainly determined by the *APOE* ⍰4 variant (rs429358-C). It is interesting to note that the *APOE* ⍰4 variant (rs429358-C) is the strongest determinant of the plasma levels of two proteins, including APOE and SNAP25. While APOE is the *cis* protein, it does not associate with MCI, tau pathology or AD in the brain, however SNAP25, which is the *trans* protein influenced by the *APOE* ⍰4 variant shows the strongest association with MCI, tau, amyloid plaques and AD in the brain. Combining the evidence from the current and previous studies, SNAP25 shows association with MCI and AD in brain, CSF and plasma and a significant MR driven by the *APOE* ⍰4 variant it is intuitive to hypothesize that *APOE* ⍰4 variant influences the risk of AD by modulating the levels of SNAP25 in plasma and brain.

From the perspective of subtyping patients for personalized medicine, an important finding of the current study is the differences observed in the proteomic signatures of AD between *APOE* ⍰4^+^ and *APOE* ⍰4^-^ prospective patients, which showed no overlap beyond GFAP and NeFL. Most of the proteins that showed significant association in the *APOE* ⍰4^-^ group did not show any association with AD in the *APOE* ⍰4^+^ group. The most interesting finding in the *APOE* ⍰4^-^ group is the association of two proteins CELSR2 and CST5 with AD. *CELSR2* hosts the genetic variant rs12740374_T, which is the primary determinant of the plasma levels of GRN^26^, and CST5 interacts with GRN through Cathepsin S (CTSS). Mutations in *GRN* are known to be causal for FTLD^30^, and a particular variant, rs5848-T in *GRN*, is believed to be causal for AD^30^. *GRN* mutations have also been associated with TAR-DNA binding protein-43 (TDP-43) pathology, which is present in 97% of ALS cases^40^, 50% of FTLD cases and up to 50% of AD cases^41^. Although no association of GRN in plasma or brain with AD has been found^30^, our findings suggest that if there is any contribution of GRN towards the development of AD, it may be in the *APOE* ⍰4^-^ AD cases. This is corroborated by our findings of the *APOE* ⍰4 stratified GWAS, where *GRN* polymorphisms show association with AD only in the *APOE* ⍰4^-^ analysis^31^.

For VaD we find statistically significant evidence of the involvement of the virus receptor pathway, which included 6 proteins, including PVR, HAVCR1, SCARB2, BSG, EPHA2 and MRC1, that were significantly associated with VaD. Of these, BSG, EPHA2, SCARB2 and HAVCR1 cluster strongly with EDA2R, which we found to be causally associated with family history of dementia and VZV seropositivity in the MR analysis but not with AD, suggesting that EDA2R is specific to vascular dementia. EDA2R (ectodysplasin A2 receptor) belongs to the tumor necrosis factor (TNF) receptor superfamily and over-expressed in the neurons^42^. In mice, EDA2R levels in the brain and plasma were shown to rise after brain injury^42^. Knocking down of EDA2R has been shown to have anti-inflammatory and antioxidant effects against lung injury^43^. The protein interaction network of EDA2R suggests involvement in multiple viral infections, including Epstein-Barr, hepatitis, measles, influenza, papillomavirus, herpes simplex 1, and SARS-Cov-1&2. These findings suggest EDA2R may be the link between viral infections and increased risk of dementia, VaD in particular. The other interesting protein is TIMP4 as it also showed a causal relationship with family history of dementia but not with AD in the MR analysis and also with VZV seropositivity. TIMP4 (tissue inhibitor of metalloproteinases 4) is over-expressed in the nervous system and heart and is known to inactivate several matrix metalloproteinases, including MMP-1, MMP-2, MMP-3, MMP-7 and MMP-9. MMP-2 and MMP-9 levels rise in the brain post ischemic injury enabling tissue remodelling and healing process^44^ but also causing breakdown the blood-brain barrier, which is associated with the initiation and progression with VaD^45^. The GWAS catalogue shows that TIMP4 is associated with vascular pathology including obesity, type 2 diabetes, hypertension and non-alcoholic fatty liver disease, and kidney function. Animal studies show involvement of TIMP4 in inflammatory processes in cardiovascular structures^46,47^, and studies in humans show upregulation of TIMP4 in cardiovascular disorders^46^. At the moment we do not have data to replicate EDA2R and TIMP4 in the brain, so their relevance to dementia remains to be explored in the brain.

Beyond the prospective causal proteins, one of the most interesting proteins is SPON1, which showed significant association with VaD in plasma, with MCI in the brain and strongest of all associations with tau and amyloid pathology but was not associated with AD, which suggests its specificity to VaD. SPON1 (spondin-1) is a cell adhesion and has been associated with vascular outcomes including obesity, atrial fibrillation, hypertension, lipoprotein measurements, infections including COVID-19 and tuberculosis, and aging-related outcomes including AD, brain connectivity and reaction time in GWAS. SPON1 is co-expressed with amyloid precursor protein (APP) and amyloid-like proteins (APLP1 & APLP2). SPON1 has been shown to inhibit APP cleavage by binding to the α/β-cleavage site of APP^48^.

We conducted the most comprehensive study to date comparing AD and VaD and exploring AD heterogeneity using plasma proteomics, validating findings in brain tissue where possible. The study has several limitations. First, UK Biobank relies on real-world clinical data, and patients with mixed AD and VaD pathology are often classified as VaD based on clinical and imaging evidence. This may increase heterogeneity within the VaD group, though we still observed distinct plasma proteomic signatures for prospective AD and VaD cases. Second, to assess causal relationships for VaD-associated proteins using MR, we used a GWAS of family history of dementia as a proxy for VaD and AD GWAS as a negative control, due to the lack of large VaD-specific GWAS. Thus, MR findings for VaD should be interpreted cautiously. Third, technical differences between plasma and brain proteomic platforms limited replication, as some plasma-identified proteins were not quantified in brain tissue. Finally, many proteins associated in the brain showed inverse associations in plasma. This may reflect biological differences—such as tissue specificity, participant age, postmortem effects, active pathology, protein turnover, or blood–brain barrier permeability^49^—or technical differences between TMT-MS, which detects post-translational modifications in brain, and affinity-based OLINK proteomics in plasma, which provides relative quantification. Similar inverse patterns across brain, CSF, and plasma have been reported in previous AD studies^49-51^.

To summarize, we identified distinct proteomic signatures of AD and VaD in plasma and confirmed these in brain. We further show that *APOE* ⍰4^-^ AD cases have a completely different proteomic signature than that of *APOE* ⍰4^+^ cases of AD, which highlights the importance of using APOE genotype in determining the course of their treatment in trials and clinical research. We identify SNAP25 as a possible causal protein for AD determined by the APOE ⍰4 genotype, suggesting that this protein may mediate the development of pathology in the brain. We identify EDA2R and TIMP4 as possible causal proteins for VaD and PVR protein as possibly causal for AD and VaD, suggesting that the vascular tissue is of interest in the mounting evidence for the role of viral infections in the development or onset of dementia.

## Data Availability

All data of the UK Biobank is available to researchers through their Research Analysis Platform (RAP).
All data of ROSMAP is available on SYNAPSE.

https://www.synapse.org/Synapse:syn3219045

https://www.ukbiobank.ac.uk/use-our-data/apply-for-access/

## Acknowledgment

We thank contributors who collected samples used in this study and patients and their families, whose help and participation made this work possible. This research has been conducted using the UK Biobank Resource under application numbers 61054. We thank the participants of ROS and MAP for their essential contributions to these projects.

## Conflicts of interest

Dr. Kaddurah-Daouk is an inventor on a series of patents on use of metabolomics for the diagnosis and treatment of CNS diseases and holds equity in Metabolon Inc., Chymia LLC and PsyProtix. Cornelia M van Duijn is currently the Research Director Brain Health of the Health Data Research UK (HDR UK) and the UK Dementia Research Institute (UK DRI), working in partnership with Dementias Platform UK (DPUK). All other authors have no disclosures to make.

## Funding

The computational aspects of this research were supported by the Wellcome Trust Core Award Grant Number 203141/Z/16/Z and the Oxford NIHR BRC. The views expressed are those of the author(s) and not necessarily those of the NHS, the NIHR or the Department of Health. ROSMAP data were provided by the Rush Alzheimer’s Disease Center, Rush University Medical Center, Chicago. Data collection was supported through funding by NIA grants P30AG10161 (ROS), R01AG15819 (ROSMAP; genomics and RNAseq), R01AG17917 (MAP), R01AG30146, R01AG36042 (5hC methylation, ATACseq), RC2AG036547 (H3K9Ac), R01AG36836 (RNAseq), R01AG48015 (monocyte RNAseq) RF1AG57473 (single nucleus RNAseq), U01AG32984 (genomic and whole exome sequencing), U01AG46152 (ROSMAP AMP-AD, targeted proteomics), U01AG46161 (TMT proteomics), U01AG61356 (whole genome sequencing, targeted proteomics, ROSMAP AMP-AD), the Illinois Department of Public Health (ROSMAP), and the Translational Genomics Research Institute (genomic). Najaf Amin is funded by National Institute on Aging (NIH) and Oxford-GSK Institute of Molecular and Computational Medicine (IMCM). Cornelia M van Duijn is supported by the US National Institute on Aging (NIH), NovoNordisk, the Oxford-GSK Institute of Molecular and Computational Medicine (IMCM), Centre of Artificial Intelligence for Precision Medicines (CAIPM) of the University of Oxford and King Abdul Aziz University, Alzheimer Research UK (ARUK), UK National Institute for Health and Care Research (NIHR) Oxford Research Center (BRC), ZonMW (Delta Dementie) and Alzheimer Nederland.

## Consent Statement

All human subjects provided informed consents.

## Notes

### Author Declarations

UK Biobank has approval from the North West Multi-Centre Research Ethics Committee, the Patient Information Advisory Group, and the Community Health Index Advisory Group. ROSMAP was approved by the Institutional Review Board (IRB) of Columbia University Medical Center (protocol AAAR4962).

## References

1. Chang Wong E, Chang Chui H. Vascular Cognitive Impairment and Dementia. Continuum (Minneap Minn). Jun 1 2022;28(3):750–780. doi:10.1212/CON.0000000000001124

2. Raulin AC, Doss SV, Trottier ZA, Ikezu TC, Bu G, Liu CC. ApoE in Alzheimer’s disease: pathophysiology and therapeutic strategies. Mol Neurodegener. Nov 8 2022;17(1):72. doi:10.1186/s13024-022-00574-4

3. Venkat P, Chopp M, Chen J. Models and mechanisms of vascular dementia. Exp Neurol. Oct 2015;272:97–108. doi:10.1016/j.expneurol.2015.05.006

4. Palmqvist S, Janelidze S, Quiroz YT, et al. Discriminative Accuracy of Plasma Phospho-tau217 for Alzheimer Disease vs Other Neurodegenerative Disorders. JAMA. Aug 25 2020;324(8):772–781. doi:10.1001/jama.2020.12134

5. Nagata K, Saito H, Ueno T, et al. Clinical diagnosis of vascular dementia. J Neurol Sci. Jun 15 2007;257(1-2):44–8. doi:10.1016/j.jns.2007.01.049

6. Lee AY. Vascular dementia. Chonnam Med J. Aug 2011;47(2):66–71. doi:10.4068/cmj.2011.47.2.66

7. Bellenguez C, Kucukali F, Jansen IE, et al. New insights into the genetic etiology of Alzheimer’s disease and related dementias. Nat Genet. Apr 2022;54(4):412–436. doi:10.1038/s41588-022-01024-z

8. Holstege H, Hulsman M, Charbonnier C, et al. Exome sequencing identifies rare damaging variants in ATP8B4 and ABCA1 as risk factors for Alzheimer’s disease. Nat Genet. Dec 2022;54(12):1786–1794. doi:10.1038/s41588-022-01208-7

9. Jansen IE, van der Lee SJ, Gomez-Fonseca D, et al. Genome-wide meta-analysis for Alzheimer’s disease cerebrospinal fluid biomarkers. Acta Neuropathol. Nov 2022;144(5):821–842. doi:10.1007/s00401-022-02454-z

10. Mega Vascular Cognitive I, Dementia c. A genome-wide association meta-analysis of all-cause and vascular dementia. Alzheimers Dement. Sep 2024;20(9):5973–5995. doi:10.1002/alz.1411511.

11. Guo Y, You J, Zhang Y, et al. Plasma proteomic profiles predict future dementia in healthy adults. Nat Aging. Feb 2024;4(2):247–260. doi:10.1038/s43587-023-00565-0

12. Ramspek CL, Steyerberg EW, Riley RD, et al. Prediction or causality? A scoping review of their conflation within current observational research. Eur J Epidemiol. Sep 2021;36(9):889–898. doi:10.1007/s10654-021-00794-w

13. Emrani S, Arain HA, DeMarshall C, Nuriel T. APOE4 is associated with cognitive and pathological heterogeneity in patients with Alzheimer’s disease: a systematic review. Alzheimers Res Ther. Nov 4 2020;12(1):141. doi:10.1186/s13195-020-00712-4

14. Shvetcov A, Johnson ECB, Winchester LM, et al. APOE epsilon4 carriers share immune-related proteomic changes across neurodegenerative diseases. Nat Med. Aug 2025;31(8):2590–2601. doi:10.1038/s41591-025-03835-z

15. Sudlow C, Gallacher J, Allen N, et al. UK biobank: an open access resource for identifying the causes of a wide range of complex diseases of middle and old age. PLoS Med. 2015/03// 2015;12(3):e1001779. doi:10.1371/journal.pmed.1001779

16. Bennett DA, Buchman AS, Boyle PA, Barnes LL, Wilson RS, Schneider JA. Religious Orders Study and Rush Memory and Aging Project. J Alzheimers Dis. 2018 2018;64(S1):S161-S189. doi:10.3233/JAD-179939

17. Vonsattel JPG, Amaya MdP, Cortes EP, Mancevska K, Keller CE. Twenty-first century brain banking: practical prerequisites and lessons from the past: the experience of New York Brain Bank, Taub Institute, Columbia University. Cell Tissue Bank. 2008/09// 2008;9(3):247–258. doi:10.1007/s10561-008-9079-y

18. Johnson ECB, Dammer EB, Duong DM, et al. Large-scale proteomic analysis of Alzheimer’s disease brain and cerebrospinal fluid reveals early changes in energy metabolism associated with microglia and astrocyte activation. Nature Medicine. 2020/05// 2020;26(5):769–780. doi:10.1038/s41591-020-0815-6

19. Biobank UK. Definitions of Dementia and the Major Diagnostic Pathologies, UK Biobank Phase 1 Outcomes Adjudication: version 1.0. March, 2018. https://biobank.ndph.ox.ac.uk/showcase/showcase/docs/alg_outcome_dementia.pdf (2018).

20. Newell KL, Hyman BT, Growdon JH, Hedley-Whyte ET. Application of the National Institute on Aging (NIA)-Reagan Institute criteria for the neuropathological diagnosis of Alzheimer disease. J Neuropathol Exp Neurol. 1999/11// 1999;58(11):1147–1155. doi:10.1097/00005072-199911000-00004

21. Braak H, Braak E. Neuropathological stageing of Alzheimer-related changes. Acta Neuropathol. 1991 1991;82(4):239–259. doi:10.1007/BF00308809

22. Wu Y, Byrne EM, Zheng Z, et al. Genome-wide association study of medication-use and associated disease in the UK Biobank. Nat Commun. 2019/04/23/ 2019;10(1):1891. doi:10.1038/s41467-019-09572-5

23. Liu J, Lahousse L, Nivard MG, et al. Integration of epidemiologic, pharmacologic, genetic and gut microbiome data in a drug-metabolite atlas. Nat Med. Jan 2020;26(1):110–117. doi:10.1038/s41591-019-0722-x

24. Bycroft C, Freeman C, Petkova D, et al. The UK Biobank resource with deep phenotyping and genomic data. Nature. Oct 2018;562(7726):203–209. doi:10.1038/s41586-018-0579-z

25. UK Biobank Genotyping of 500,000 UK Biobank participants. version 2.0. https://biobank.ndph.ox.ac.uk/showcase/showcase/docs/ukb_dna_processing.pdf (2017).

26. Sun BB, Chiou J, Traylor M, et al. Plasma proteomic associations with genetics and health in the UK Biobank. Nature. Oct 2023;622(7982):329–338. doi:10.1038/s41586-023-06592-6

27. Zhang Z, Gayle AA, Wang J, Zhang H, Cardinal-Fernandez P. Comparing baseline characteristics between groups: an introduction to the CBCgrps package. Ann Transl Med. Dec 2017;5(24):484. doi:10.21037/atm.2017.09.39

28. Kunkle BW, Grenier-Boley B, Sims R, et al. Genetic meta-analysis of diagnosed Alzheimer’s disease identifies new risk loci and implicates Abeta, tau, immunity and lipid processing. Nat Genet. Mar 2019;51(3):414–430. doi:10.1038/s41588-019-0358-2

29. Rhinn H, Tatton N, McCaughey S, Kurnellas M, Rosenthal A. Progranulin as a therapeutic target in neurodegenerative diseases. Trends Pharmacol Sci. Aug 2022;43(8):641–652. doi:10.1016/j.tips.2021.11.015

30. Wang XM, Zeng P, Fang YY, Zhang T, Tian Q. Progranulin in neurodegenerative dementia. J Neurochem. Jul 2021;158(2):119–137. doi:10.1111/jnc.15378

31. Thomassen JQ, Hampton L, Ulms B, et al. APOE stratified genome-wide association studies provide novel insights into the genetic etiology of Alzheimers’s disease. medRxiv. 2025:2025.05.07.25327065. doi:10.1101/2025.05.07.25327065

32. Wightman DP, Jansen IE, Savage JE, et al. A genome-wide association study with 1,126,563 individuals identifies new risk loci for Alzheimer’s disease. Nat Genet. Sep 2021;53(9):1276–1282. doi:10.1038/s41588-021-00921-z

33. Blackhurst BM, Funk KE. Viral pathogens increase risk of neurodegenerative disease. Nat Rev Neurol. May 2023;19(5):259–260. doi:10.1038/s41582-023-00790-6

34. Halbgebauer S, Steinacker P, Hengge S, et al. CSF levels of SNAP-25 are increased early in Creutzfeldt-Jakob and Alzheimer’s disease. J Neurol Neurosurg Psychiatry. Aug 22 2022;doi:10.1136/jnnp-2021-328646

35. Wingo AP, Liu Y, Gerasimov ES, et al. Integrating human brain proteomes with genome-wide association data implicates new proteins in Alzheimer’s disease pathogenesis. Nat Genet. Feb 2021;53(2):143–146. doi:10.1038/s41588-020-00773-z

36. Antonucci F, Corradini I, Fossati G, Tomasoni R, Menna E, Matteoli M. SNAP-25, a Known Presynaptic Protein with Emerging Postsynaptic Functions. Front Synaptic Neurosci. 2016;8:7. doi:10.3389/fnsyn.2016.00007

37. Oyler GA, Higgins GA, Hart RA, et al. The identification of a novel synaptosomal-associated protein, SNAP-25, differentially expressed by neuronal subpopulations. J Cell Biol. Dec 1989;109(6 Pt 1):3039–52. doi:10.1083/jcb.109.6.3039

38. Gosso MF, de Geus EJ, van Belzen MJ, et al. The SNAP-25 gene is associated with cognitive ability: evidence from a family-based study in two independent Dutch cohorts. Mol Psychiatry. Sep 2006;11(9):878–86. doi:10.1038/sj.mp.4001868

39. Guerini FR, Agliardi C, Sironi M, et al. Possible association between SNAP-25 single nucleotide polymorphisms and alterations of categorical fluency and functional MRI parameters in Alzheimer’s disease. J Alzheimers Dis. 2014;42(3):1015–28. doi:10.3233/JAD-140057

40. Scotter EL, Chen HJ, Shaw CE. TDP-43 Proteinopathy and ALS: Insights into Disease Mechanisms and Therapeutic Targets. Neurotherapeutics. Apr 2015;12(2):352–63. doi:10.1007/s13311-015-0338-x

41. Jo M, Lee S, Jeon YM, Kim S, Kwon Y, Kim HJ. The role of TDP-43 propagation in neurodegenerative diseases: integrating insights from clinical and experimental studies. Exp Mol Med. Oct 2020;52(10):1652–1662. doi:10.1038/s12276-020-00513-7

42. Lastra Romero A, Seitz T, Zisiadis GA, Jeffery H, Osman AM. EDA2R reflects the acute brain response to cranial irradiation in liquid biopsies. Neuro-Oncology. 2024;26(9):1617–1627. doi:10.1093/neuonc/noae077

43. Jia N, Jia Y, Yang F, Du W. Knockdown of EDA2R alleviates hyperoxia-induced lung epithelial cell injury by inhibiting NF-kappaB pathway. Allergol Immunopathol (Madr). 2022;50(5):84–90. doi:10.15586/aei.v50i5.670

44. Wysocka A, Szczygielski J, Kopanska M, Oertel JM, Glowniak A. Matrix Metalloproteinases in Cardioembolic Stroke: From Background to Complications. Int J Mol Sci. Feb 11 2023;24(4)doi:10.3390/ijms24043628

45. Du SQ, Wang XR, Xiao LY, et al. Molecular Mechanisms of Vascular Dementia: What Can Be Learned from Animal Models of Chronic Cerebral Hypoperfusion? Mol Neurobiol. Jul 2017;54(5):3670–3682. doi:10.1007/s12035-016-9915-1

46. Koskivirta I, Rahkonen O, Mayranpaa M, et al. Tissue inhibitor of metalloproteinases 4 (TIMP4) is involved in inflammatory processes of human cardiovascular pathology. Histochem Cell Biol. Sep 2006;126(3):335–42. doi:10.1007/s00418-006-0163-8

47. Schulze CJ, Wang W, Suarez-Pinzon WL, Sawicka J, Sawicki G, Schulz R. Imbalance between tissue inhibitor of metalloproteinase-4 and matrix metalloproteinases during acute myocardial [correction of myoctardial] ischemia-reperfusion injury. Circulation. May 20 2003;107(19):2487–92. doi:10.1161/01.CIR.0000065603.09430.58

48. Ho A, Sudhof TC. Binding of F-spondin to amyloid-beta precursor protein: a candidate amyloid-beta precursor protein ligand that modulates amyloid-beta precursor protein cleavage. Proc Natl Acad Sci U S A. Feb 24 2004;101(8):2548–53. doi:10.1073/pnas.0308655100

49. Farinas A, Rutledge J, Bot VA, et al. Disruption of the cerebrospinal fluid-plasma protein balance in cognitive impairment and aging. Nat Med. Jul 15 2025;doi:10.1038/s41591-025-03831-350.

50. Guo Q, Ping L, Dammer EB, et al. Heparin-enriched plasma proteome is significantly altered in Alzheimer’s disease. Mol Neurodegener. Oct 8 2024;19(1):67. doi:10.1186/s13024-024-00757-151.

51. Spies PE, Verbeek MM, van Groen T, Claassen JA. Reviewing reasons for the decreased CSF Abeta42 concentration in Alzheimer disease. Front Biosci (Landmark Ed). Jun 1 2012;17(6):2024–34. doi:10.2741/4035

