## Supplementary Figures for "Proteomic profiling of Alzheimer’s disease and Vascular dementia reveals unique underlying signatures"

**Supplementary Figure Legends**

**Supplementary Figure 1a**: Correlation structure of 55 proteins associated with AD in cases.

**Supplementary Figure 1b**: Correlation structure of 55 proteins associated with AD in controls.

**Supplementary Figure 2**: Overlap in the AD associated proteins in *APOE Ɛ*4^+^ and *APOE Ɛ*4^-^ carriers and overall.

**Supplementary Figure 3**: Genetic variants in CELSR2 locus determine plasma levels of progranulin (GRN).

**Supplementary Figure 4**: Interaction network of CST5.

**Supplementary Figure 5**: Scatterplot of proteomic signatures of AD and VaD. Axes depict the Z-scores from proteome-wide association analyses in the two strata. Each diamond represents a protein, where FDR significant ones are highlighted in blue, red and black. Blue are the proteins uniquely associated with VaD and red are the proteins uniquely associated with AD. Black diamonds are the proteins associated with both types of dementia.

**Supplementary Figure 6a**: Correlation structure of the 49 proteins associated with VaD in cases.

**Supplementary Figure 6b**: Correlation structure of the 49 proteins associated with VaD in controls.

**Supplementary Figure 7**: Hierarchical clustering of dementias based on proteomic signatures.

**Supplementary Figure 8:** Forest plots and Results of leave-one-out analysis for APOE, SNAP25 and PALM.

**Supplementary Figure 9:** Interaction network of APOE, SNAP25 and PALM

**Supplementary Figure 10:** PVR is an independent locus in AD GWAS near the APOE region

**Supplementary Figure 11:** Forest plot and leave-one-out analysis with AD as exposure and plasma SNAP25 levels as the outcome in MR.

**Supplementary Figure 12**: Leave-one-out results with AD and FHAD as exposures and plasma PVR levels as outcome in MR.

**Supplementary Figure 1 a: Correlation structure of 55 proteins associated with AD in cases**

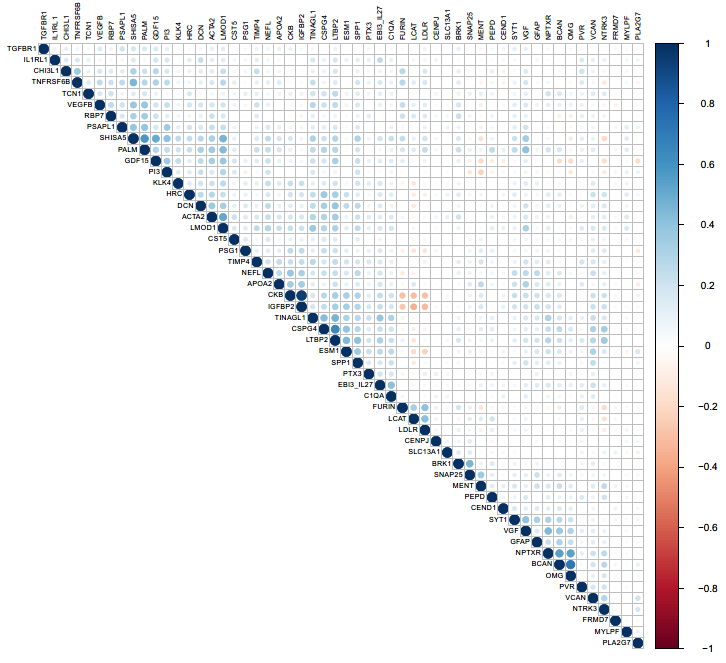

**Supplementary Figure 1 b: Correlation structure of 55 proteins associated with AD in controls**

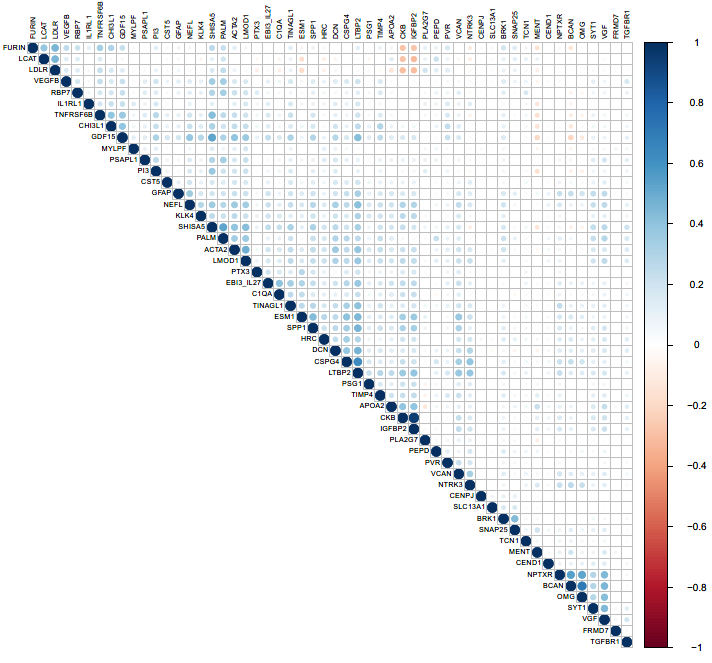

**Supplementary Figure 2: Overlap in the AD associated proteins in *APOE Ɛ*4^+^ and *APOE Ɛ*4^-^ carriers and overall.**

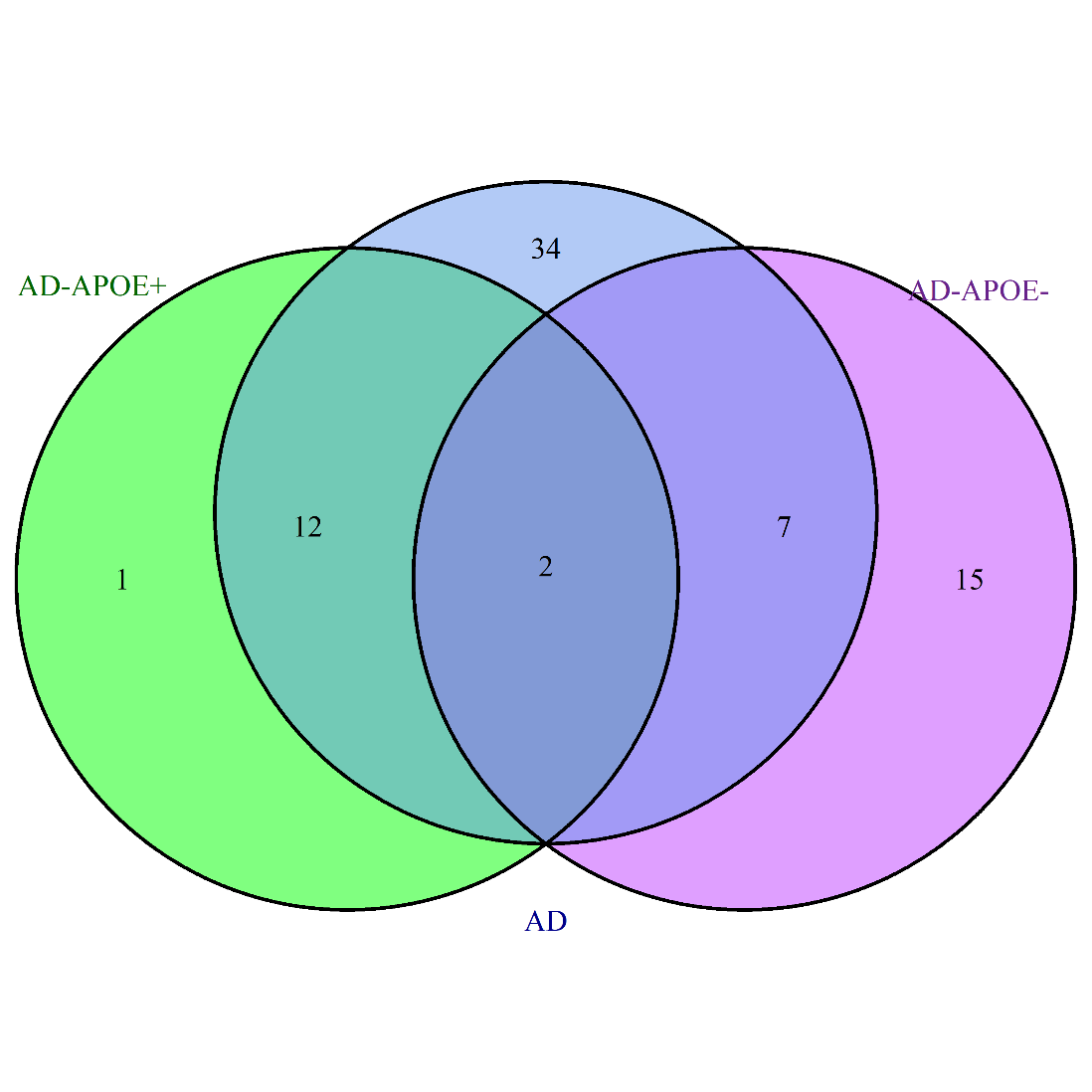

LTBP2 SHISA5 CST5 VEGFB LMOD TGFBR1 FRMD7

NPTX2 ADGRD1

DTX3 BMPER

ALSTN2

GFRA2

FLT1

KLK13

IL24

YAP1 CTHRC1 FNLN2

GUCY2C

SMOC2

CELSR2

VGF

SYT1 CEND1 MENT NPTXR LDLR IL1RL1 RBP7

EBI3-IL27

TNFRSF6B SLC13A1

PSG1

STAB2

GFAP

NEFL

IGFBP2, PTX3, PEPD, BRK1, CENPJ, ESM1, APOE, OMG, MYLPF, KLK4, PVR, DCN, BCAN, CKB, TINAGL1, SPP1, CHI3L1, PI3, GDF15, TIMP4, C1QA, TCN1, VCAN, LCAT, FURIN, CSPG4, SNAP25, ACTA2, PSAPL1, APOA2, NTRK3, HRC, PLA2G7, PALM

**Supplementary Figure 3: Genetic variants in CELSR2 locus determine plasma levels of progranulin (GRN).**

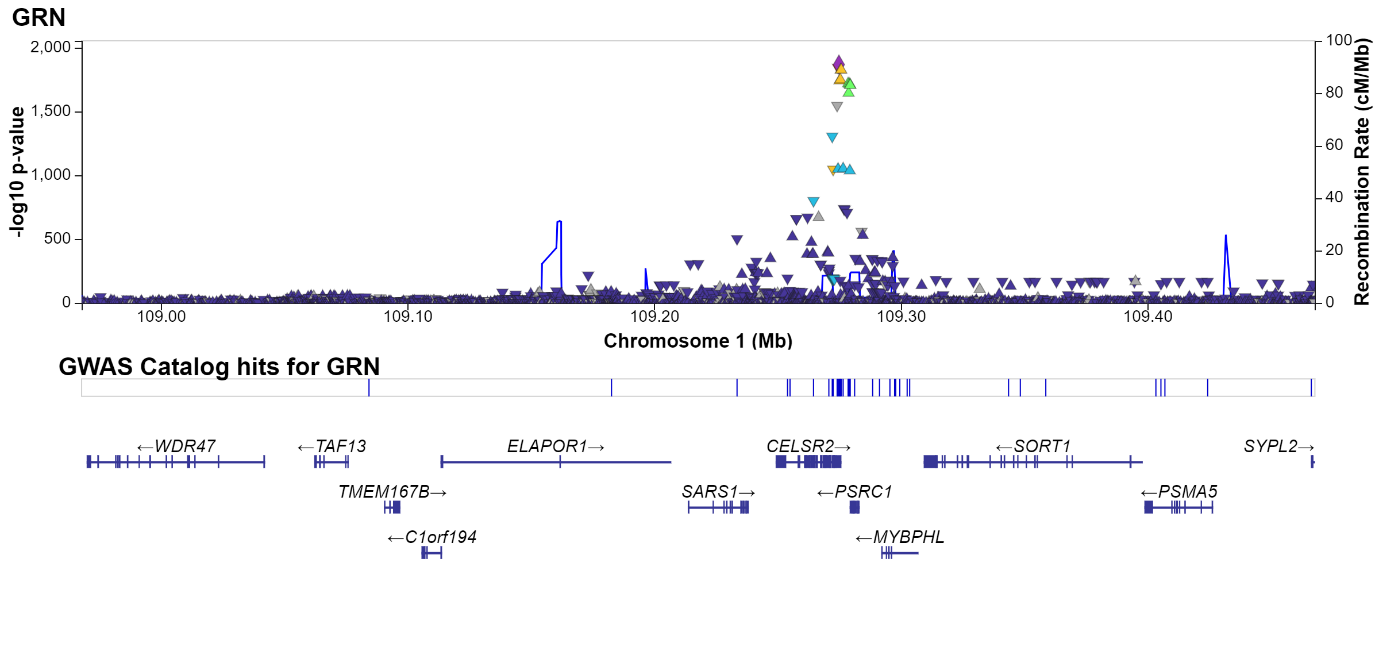

**Supplementary Figure 4: Interaction network of CST5.**

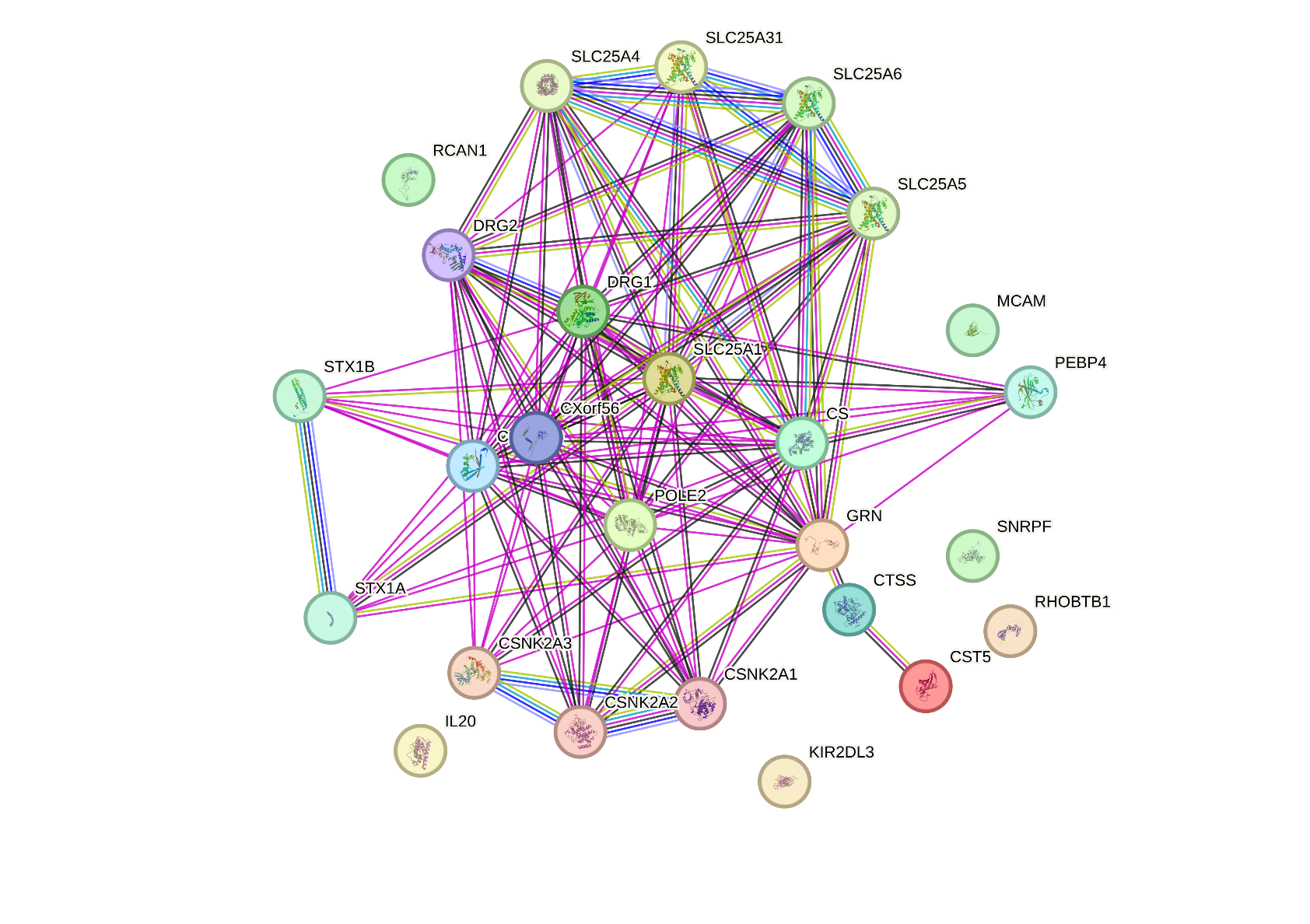

**Supplementary Figure 5: Scatterplot of proteomic signatures of AD and VAD**

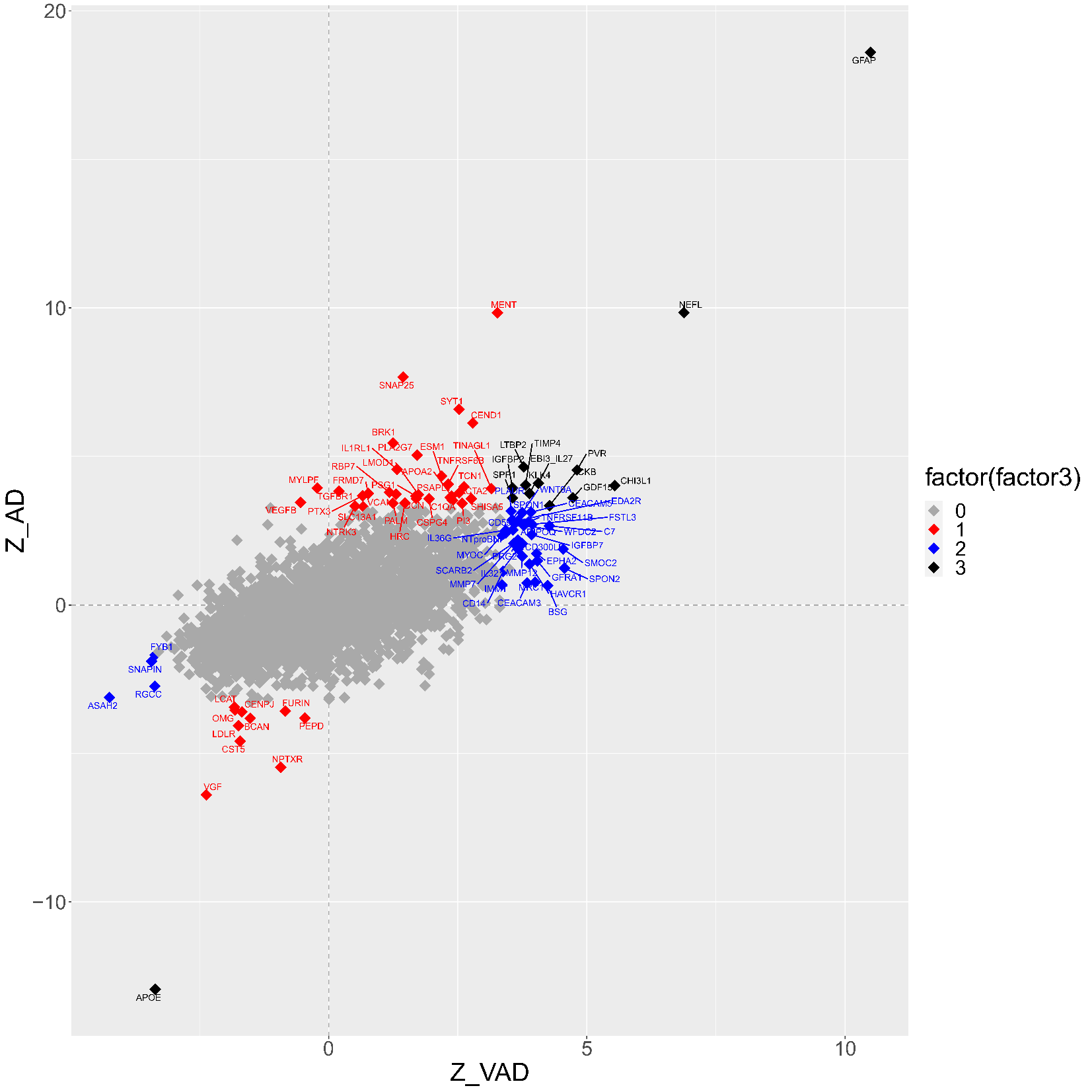

**Supplementary Figure 6a: Correlation structure of the 49 proteins associated with VAD in cases**

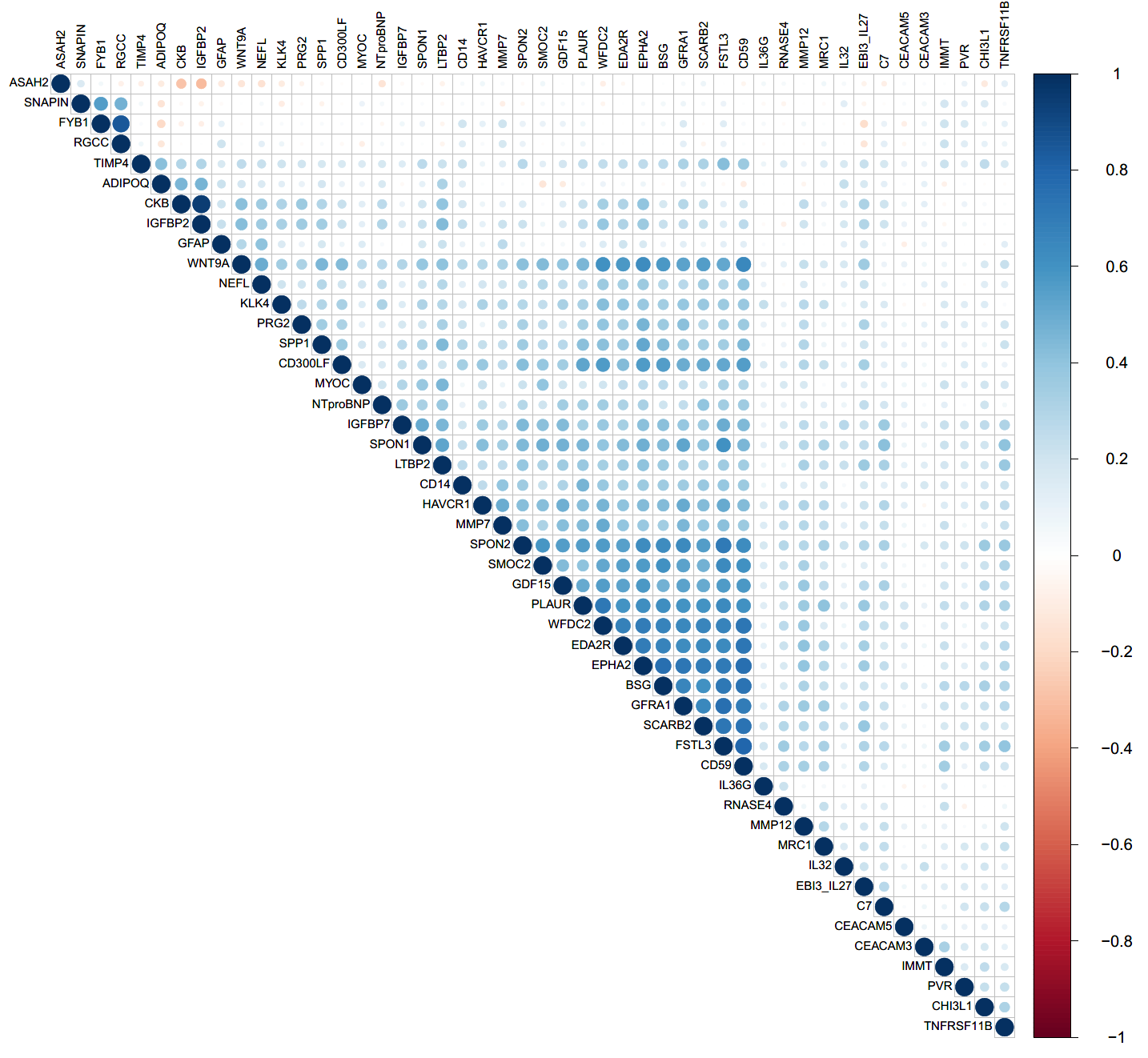

**Supplementary Figure 6b: Correlation structure of the 49 proteins associated with VAD in controls**

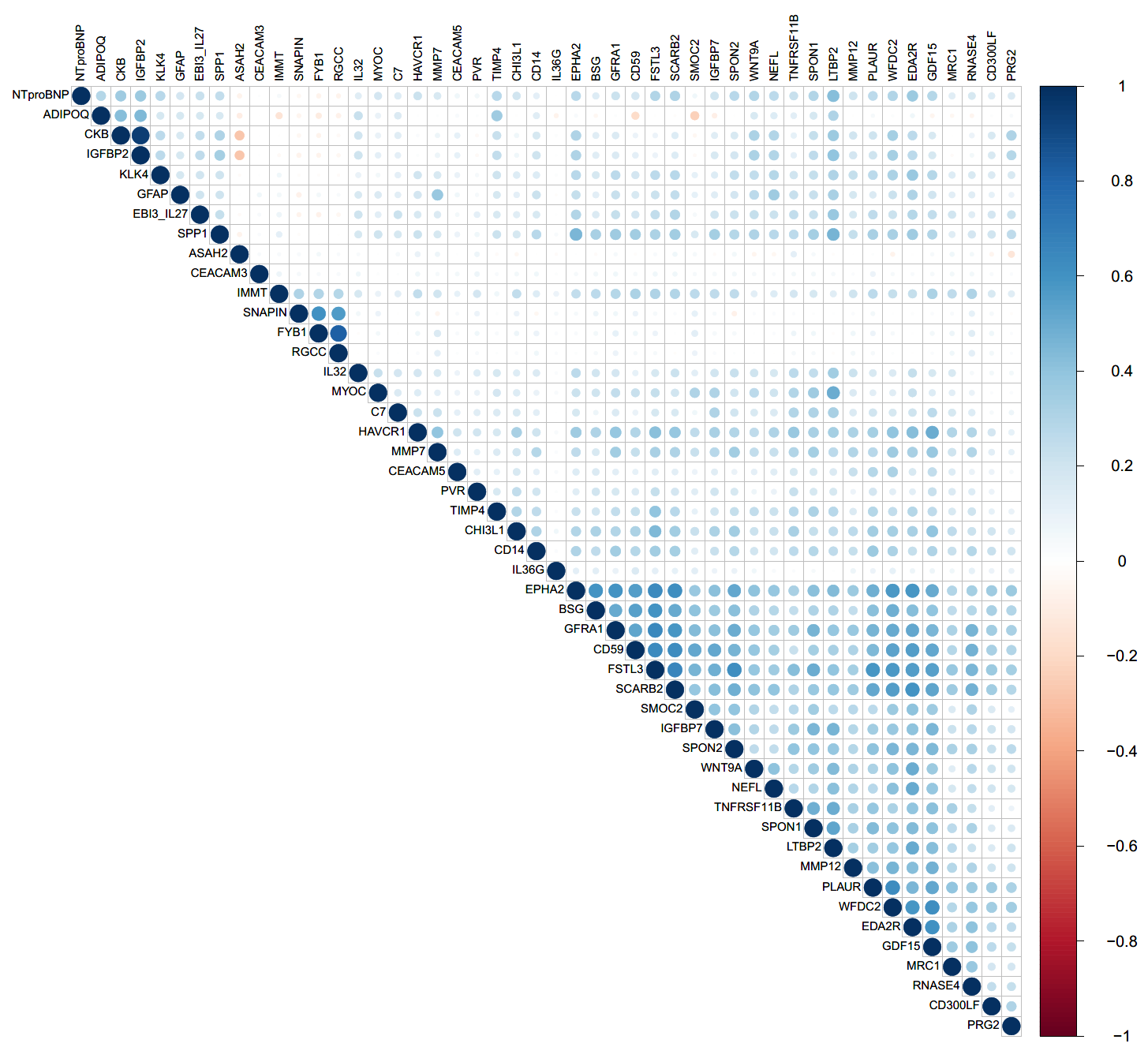

**Supplementary Figure 7: Hierarchical clustering of dementias based on proteomic signatures**

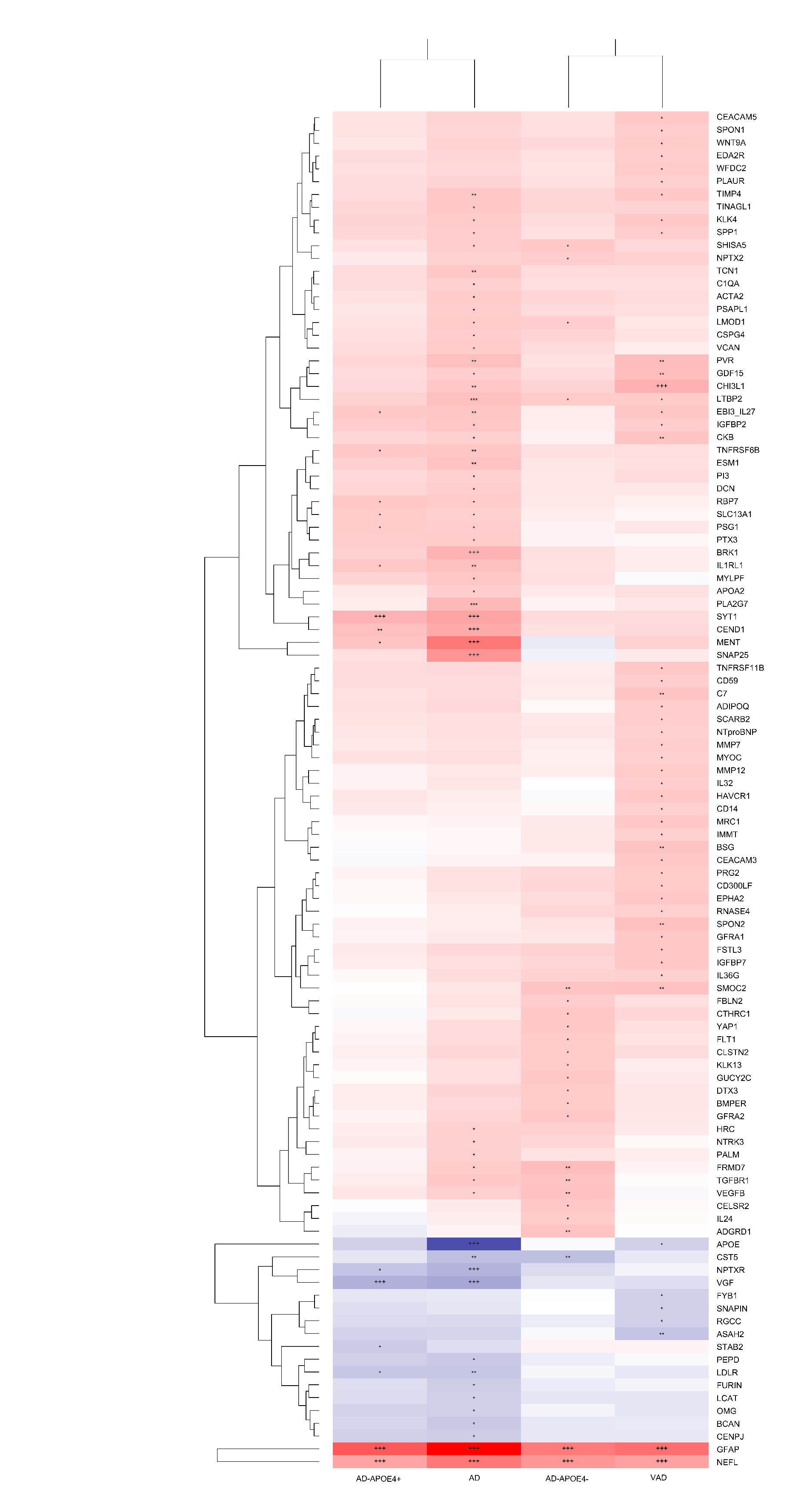

**Supplementary Figure 8: Forest plots and Results of leave-one-out analysis for APOE, SNAP25 and PALM**

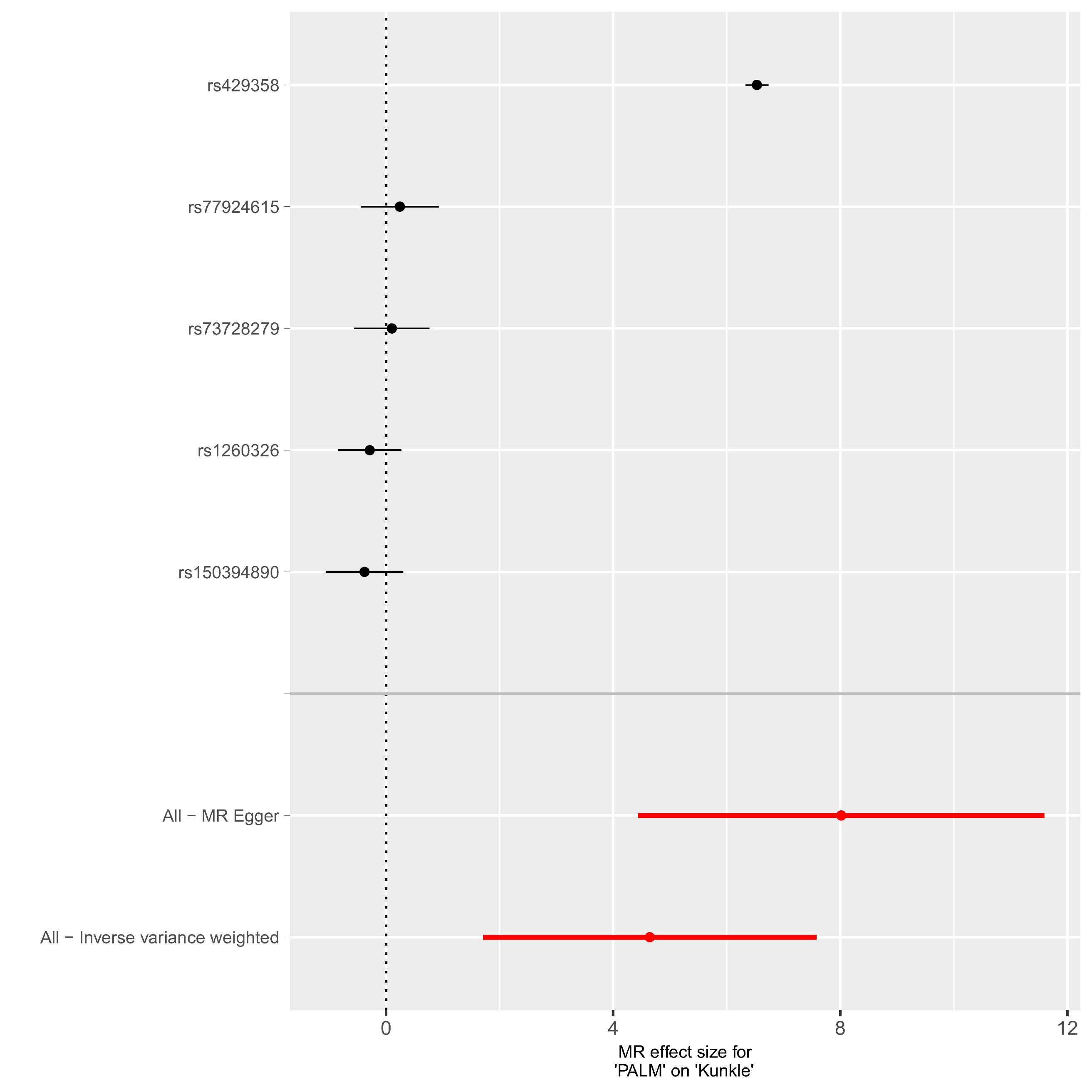

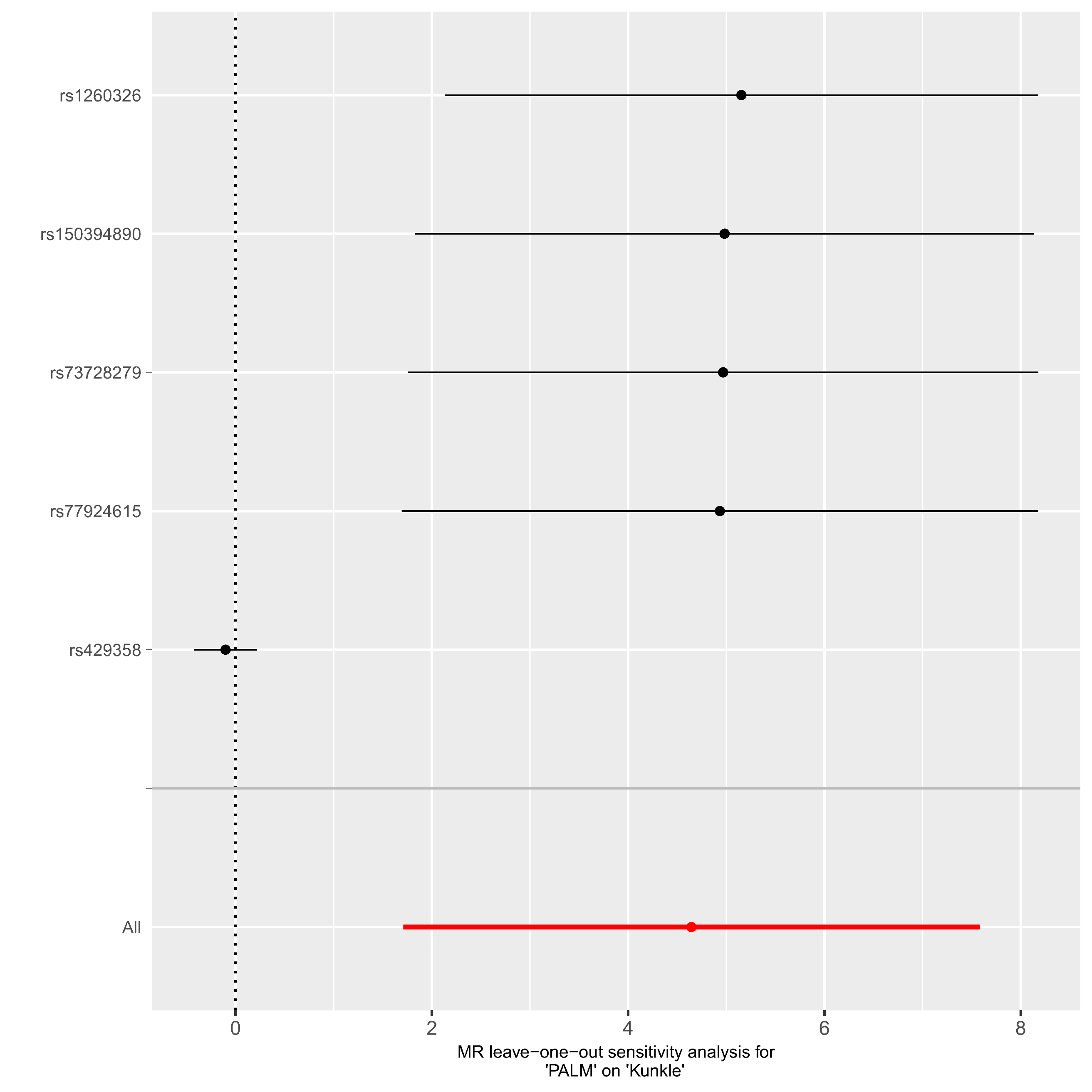

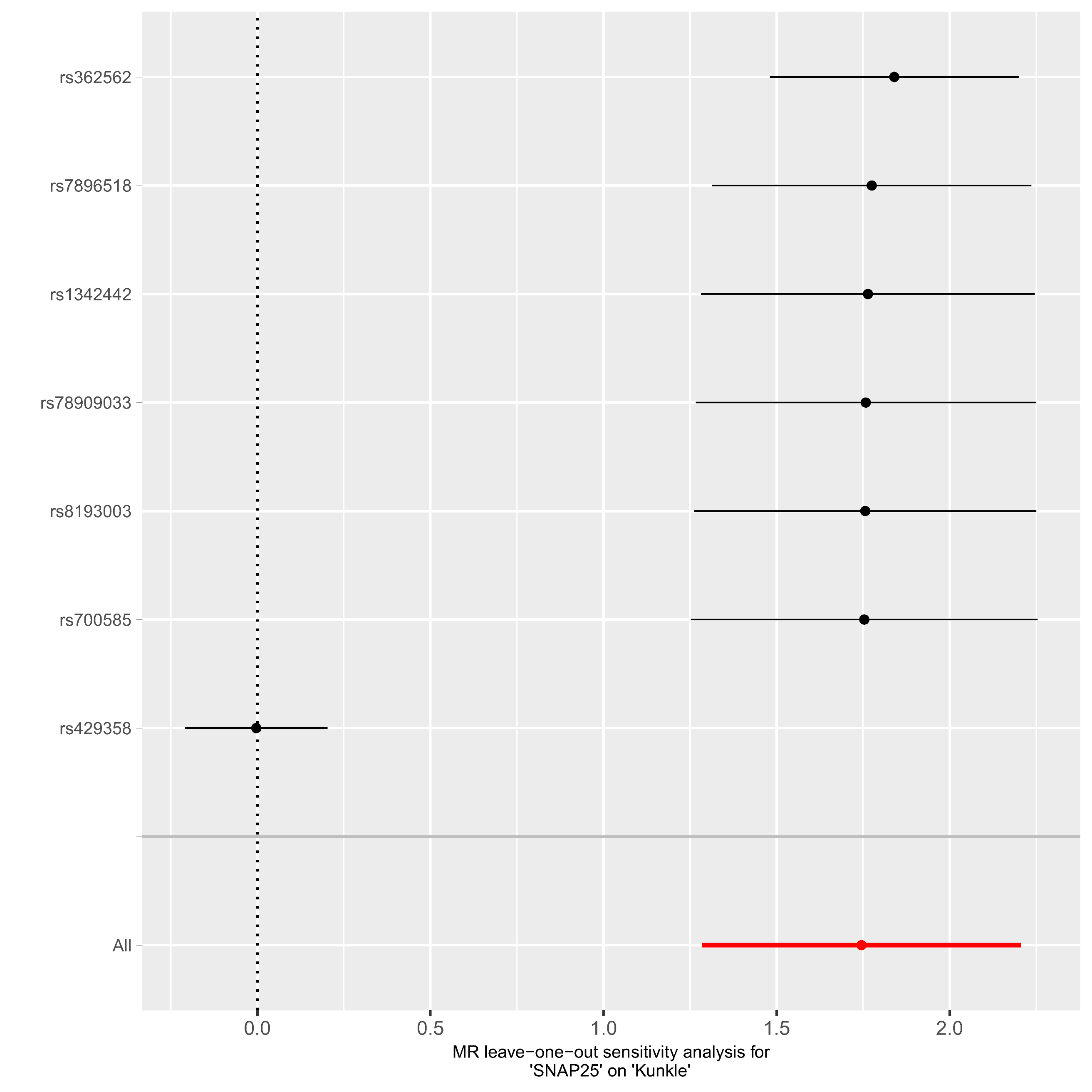

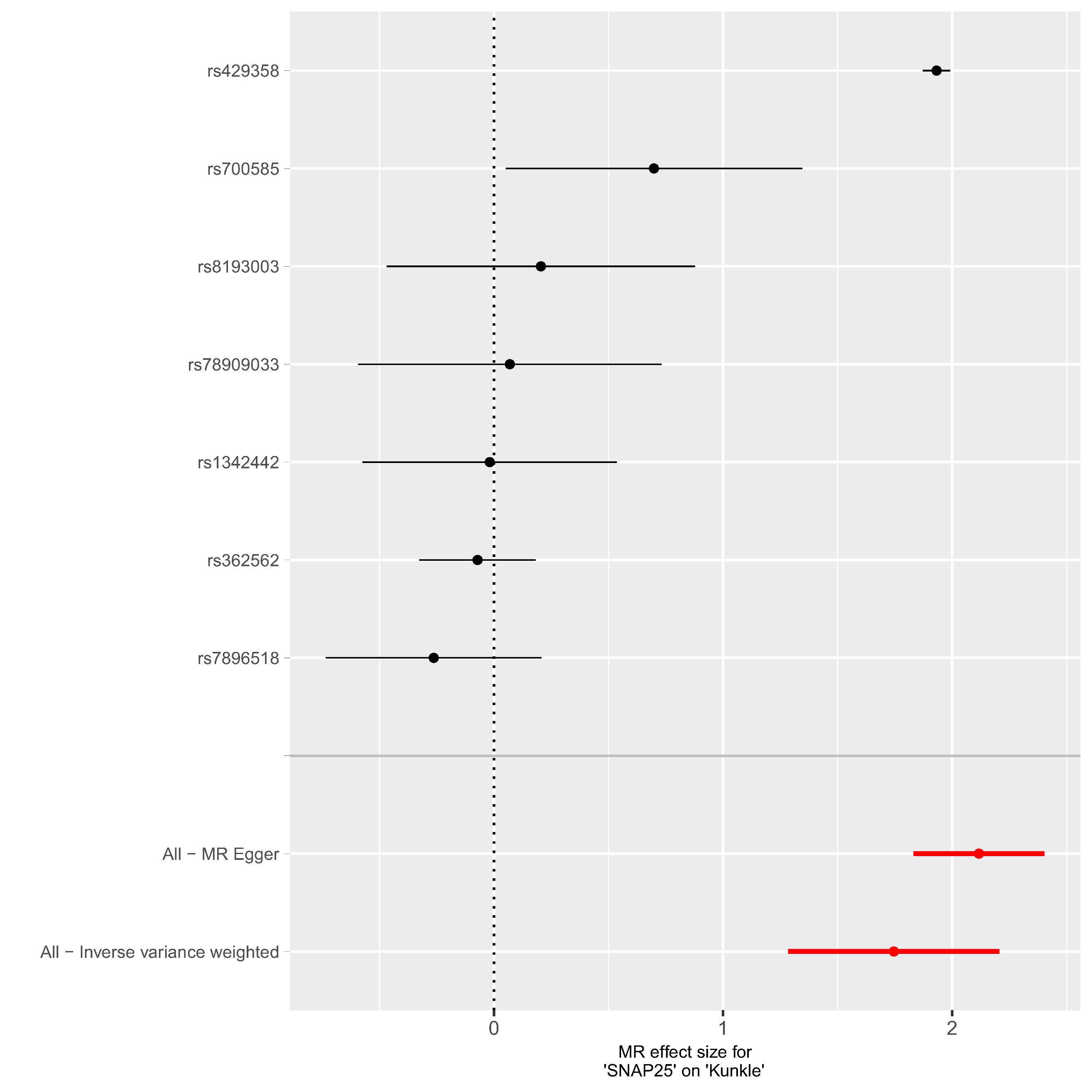

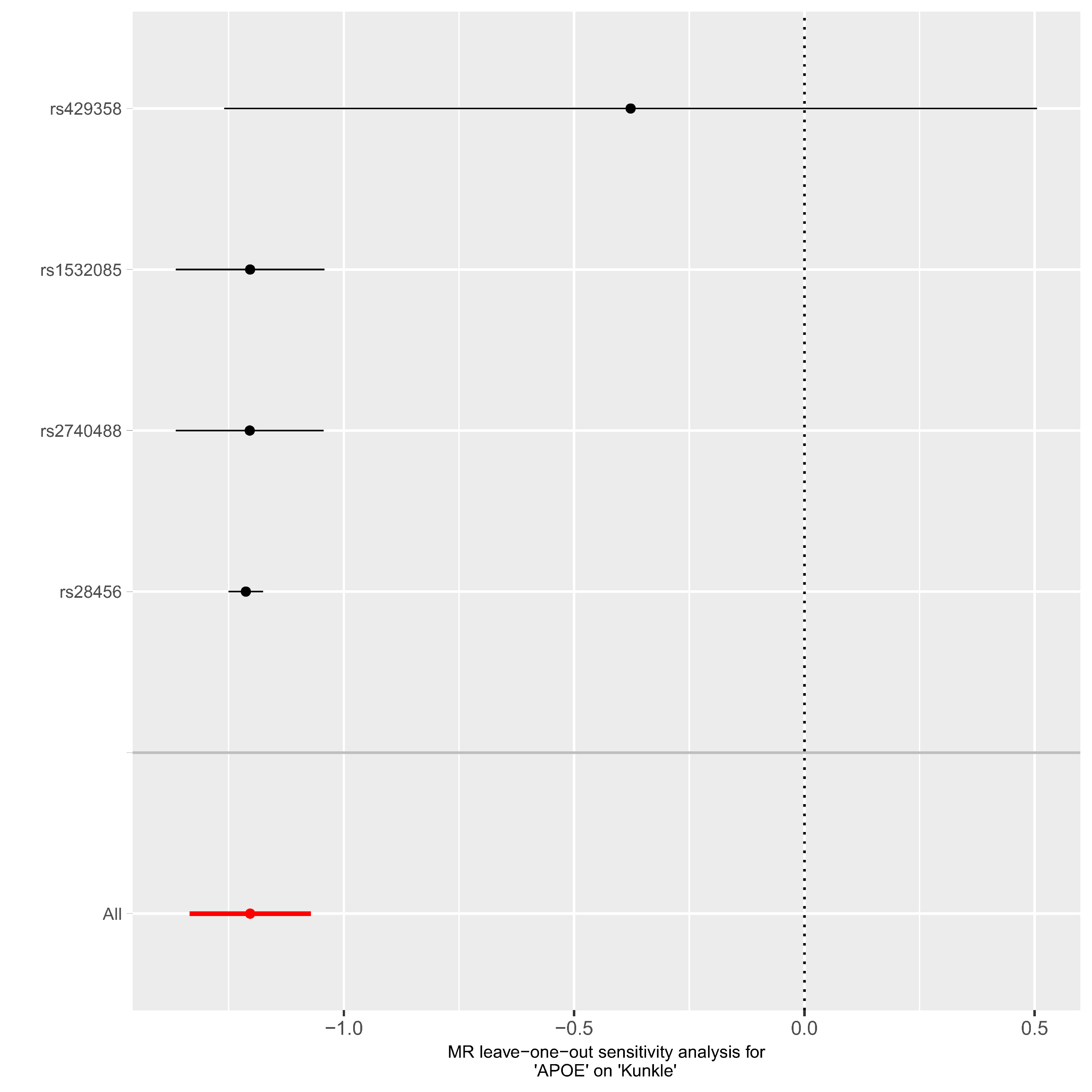

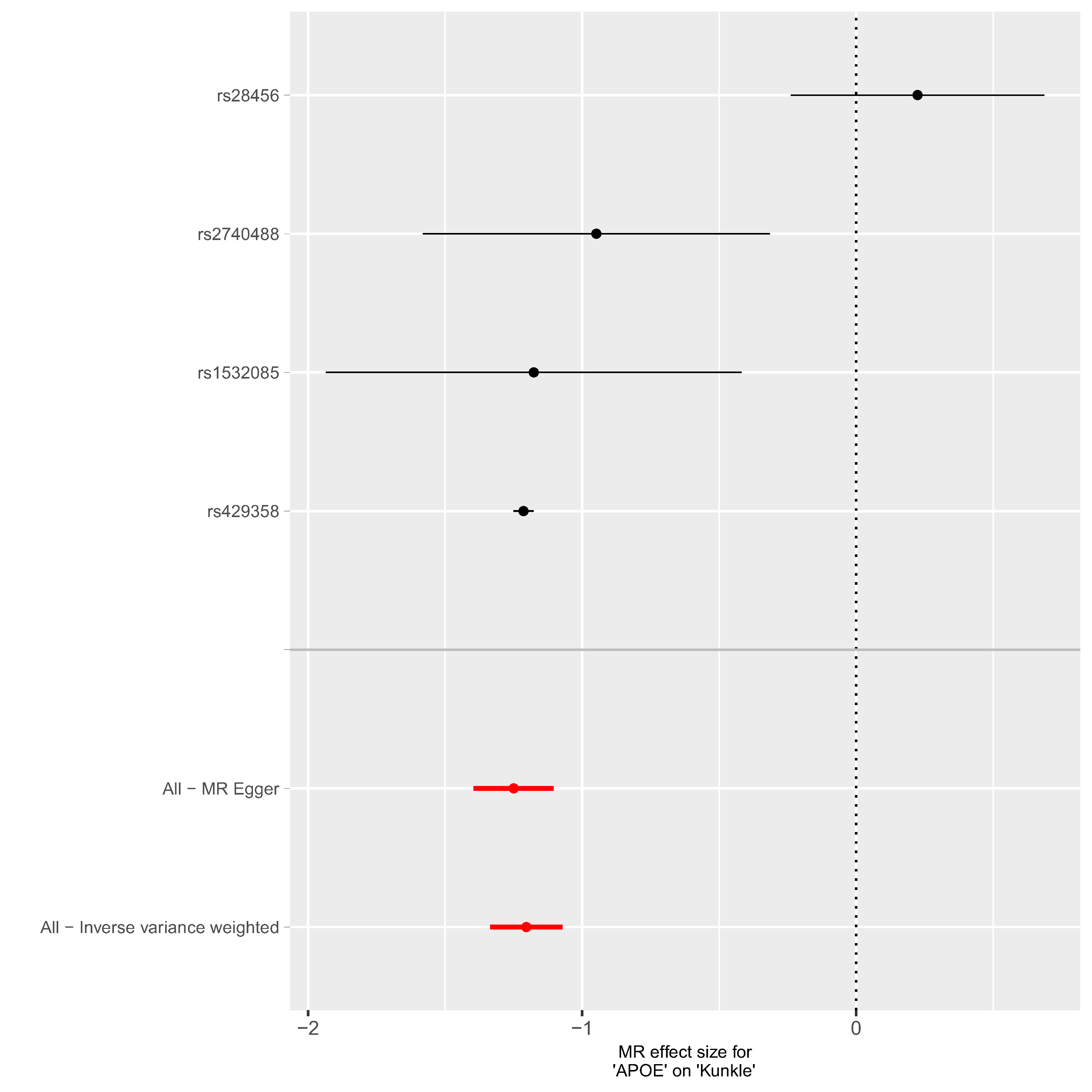

**Supplementary Figure 9: Interaction network of APOE, SNAP25 and PALM**

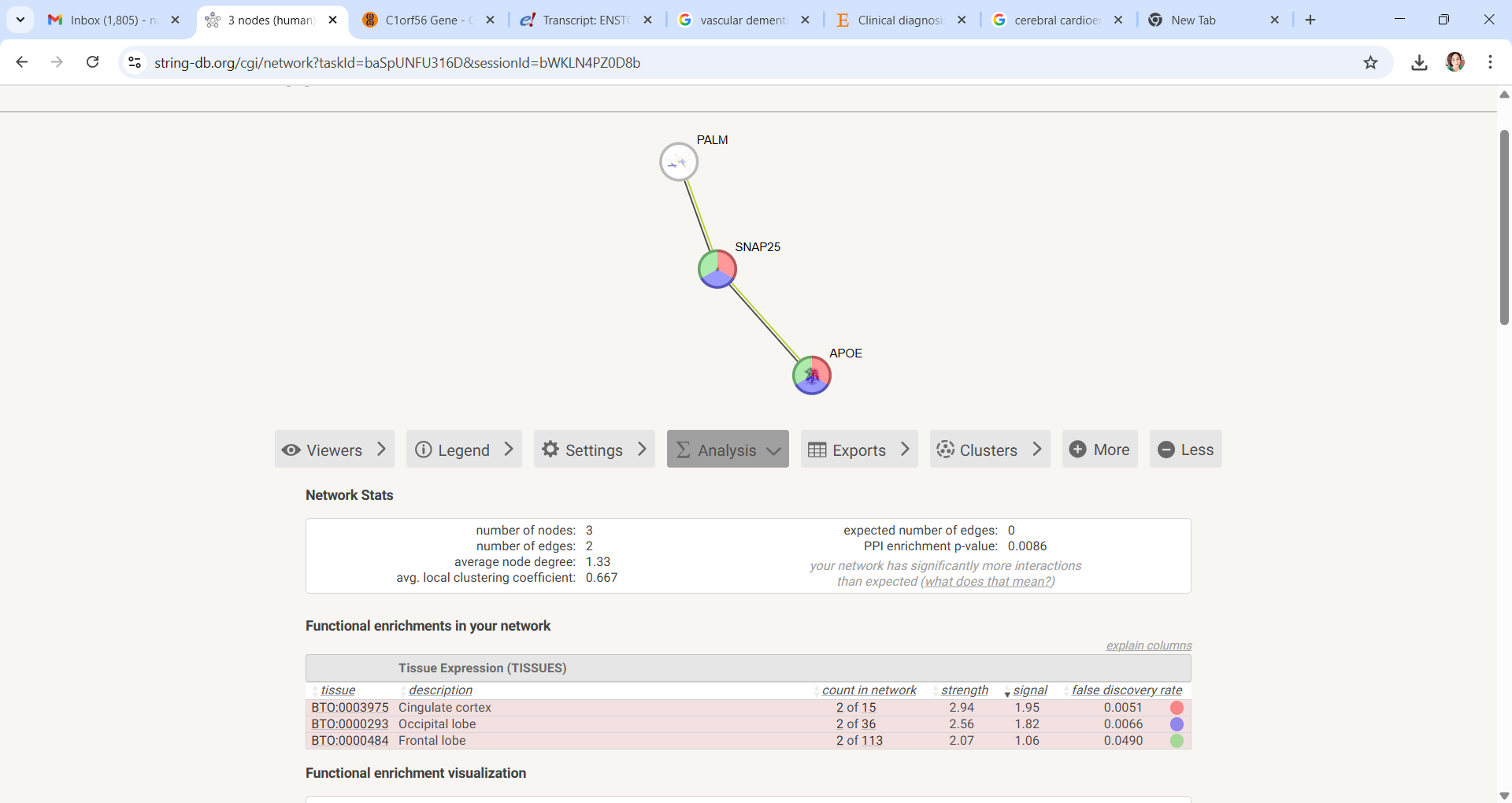

Supplementary Figure 10: PVR is an independent locus in AD GWAS near the APOE region

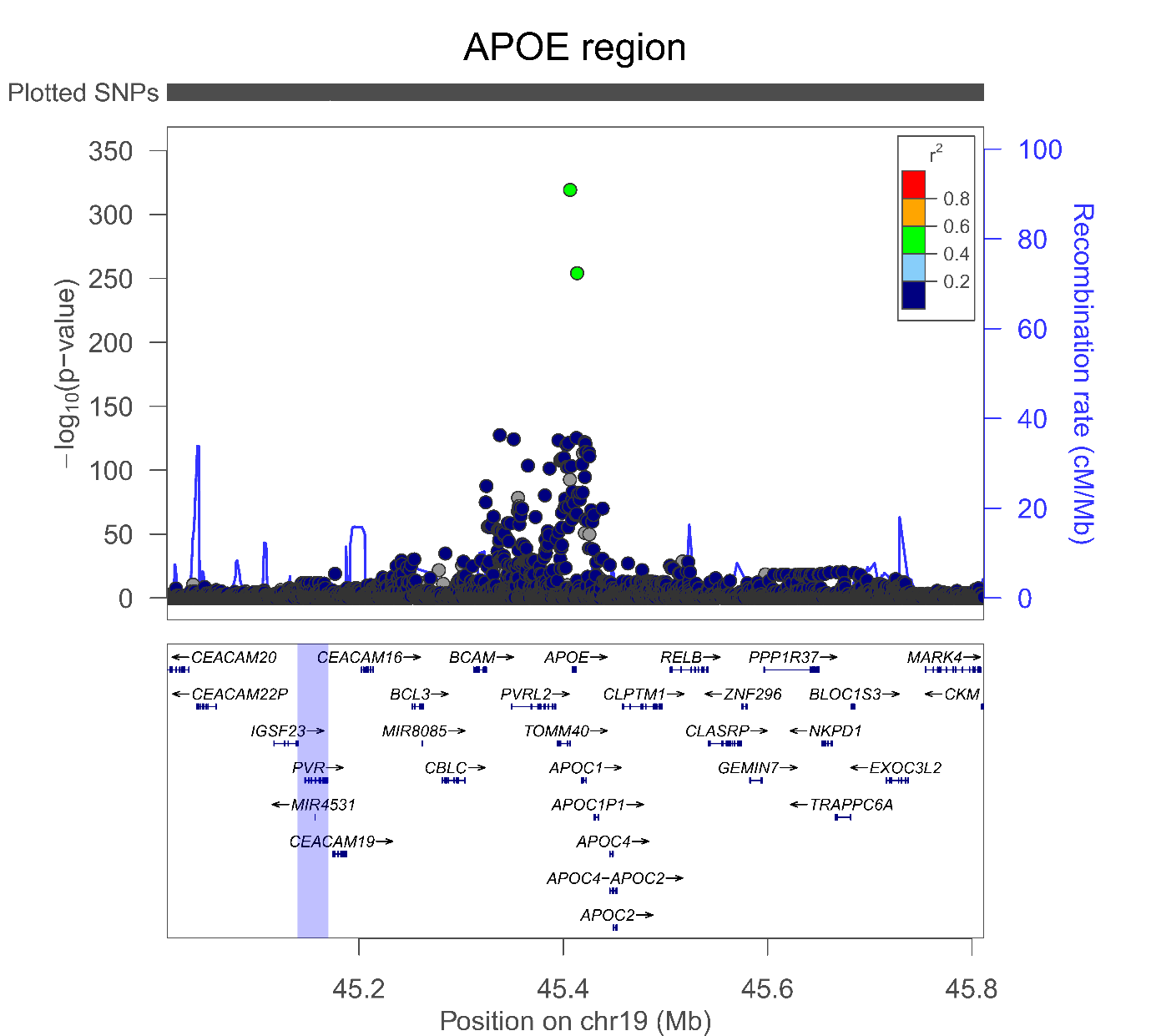

**Supplementary Figure 11: Forest plot and leave-one-out analysis with AD as exposure and plasma SNAP25 levels as the outcome in MR.**

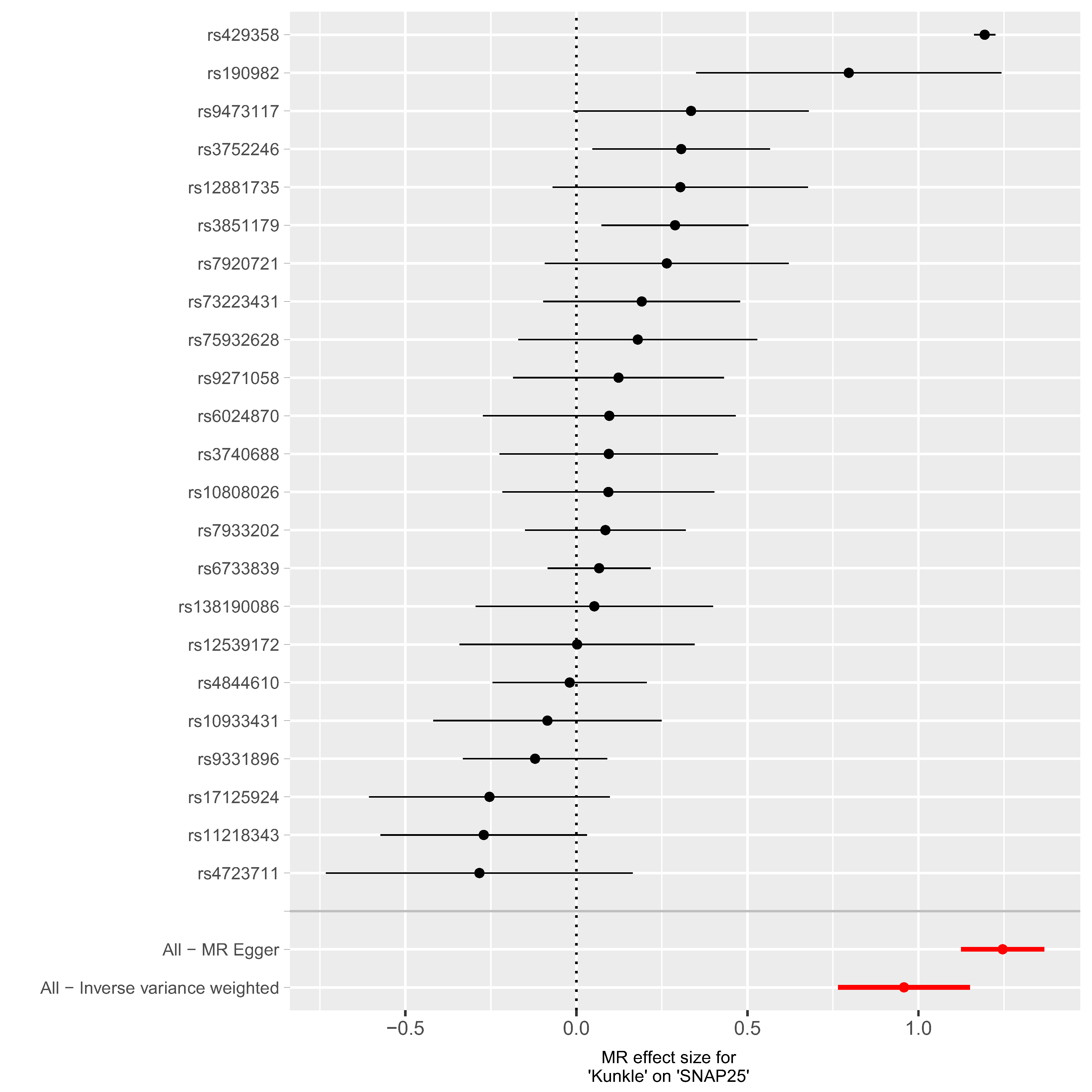

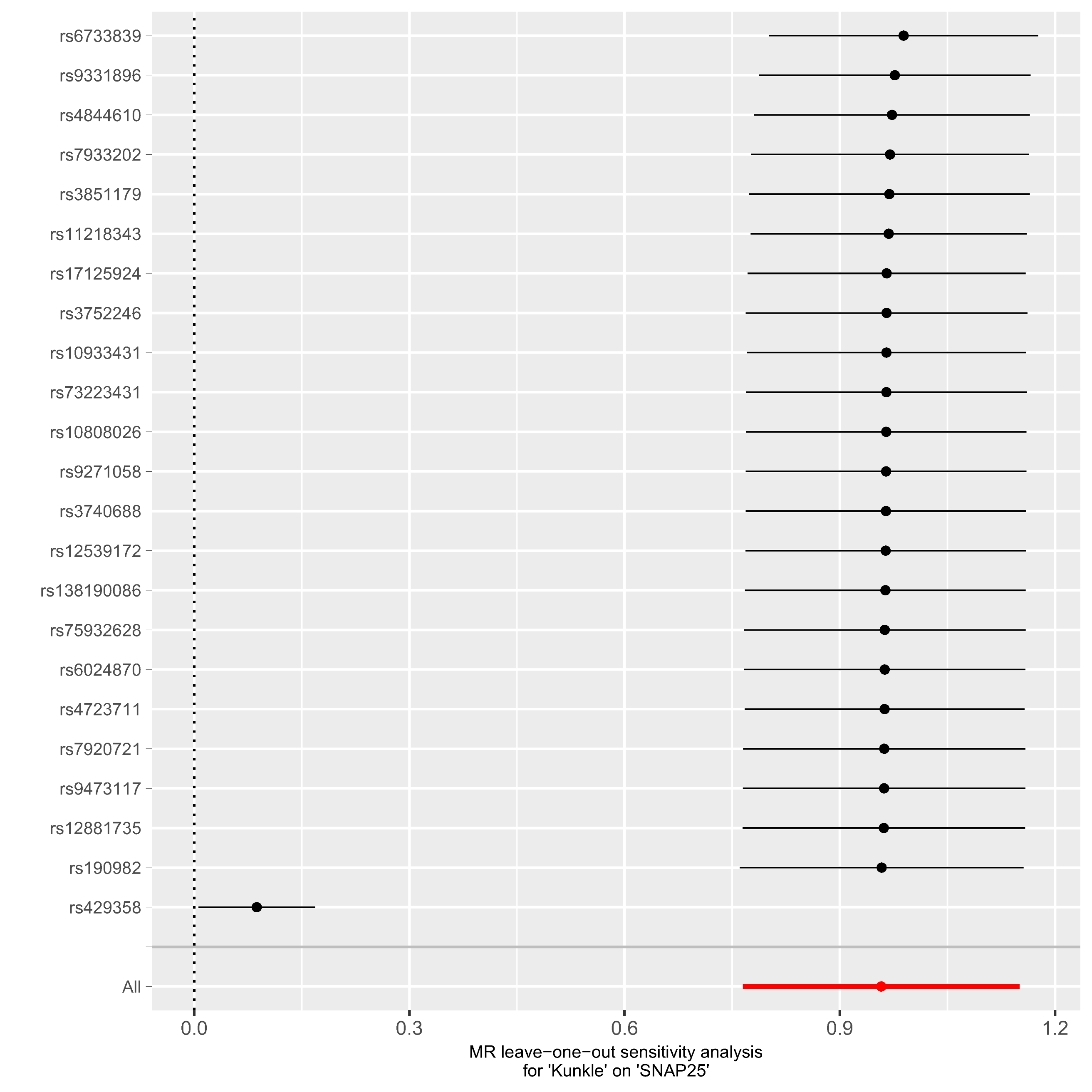

Supplementary Figure 12: Leave-one-out results with AD and FHAD as exposures and plasma PVR levels as outcome in MR.

AD

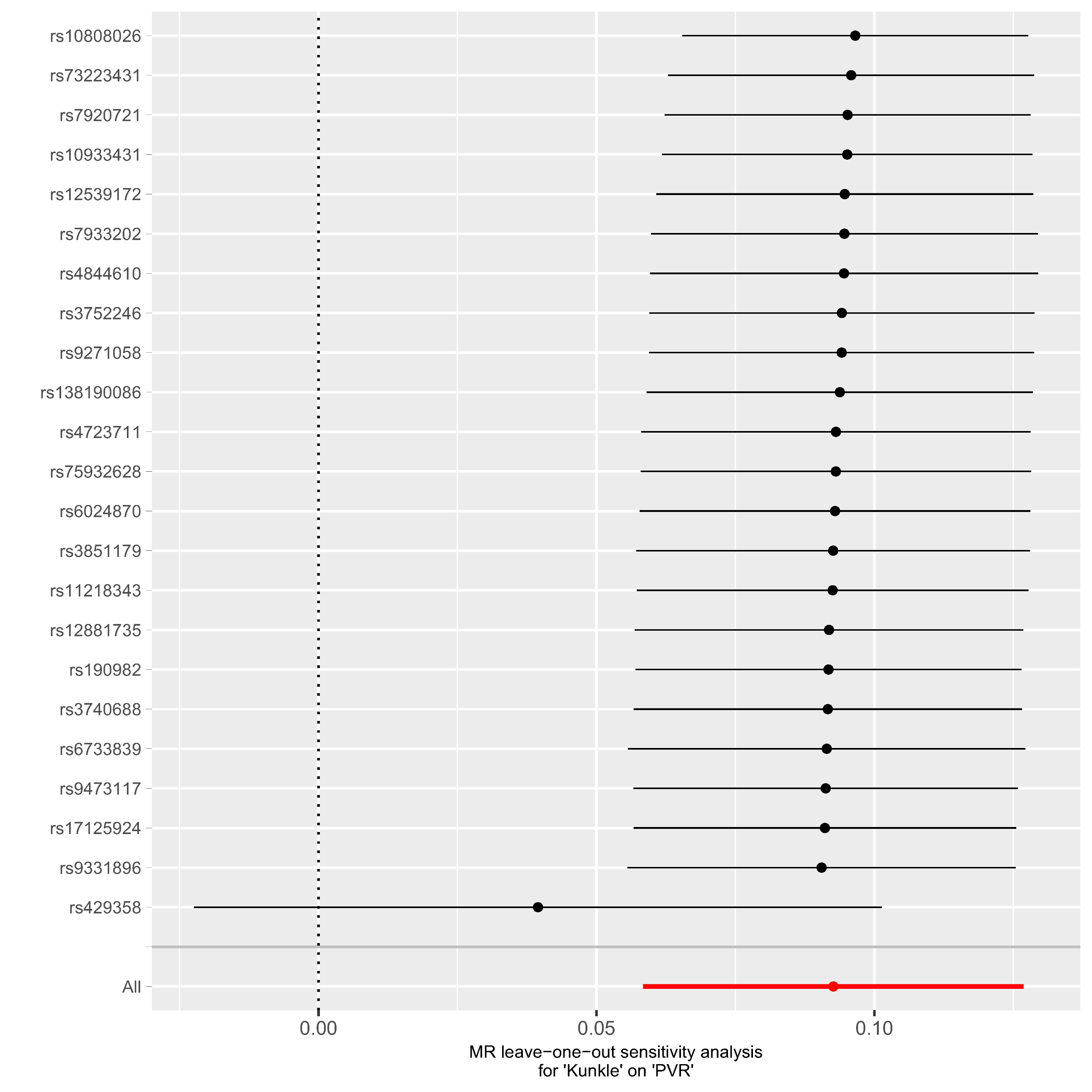

FHAD

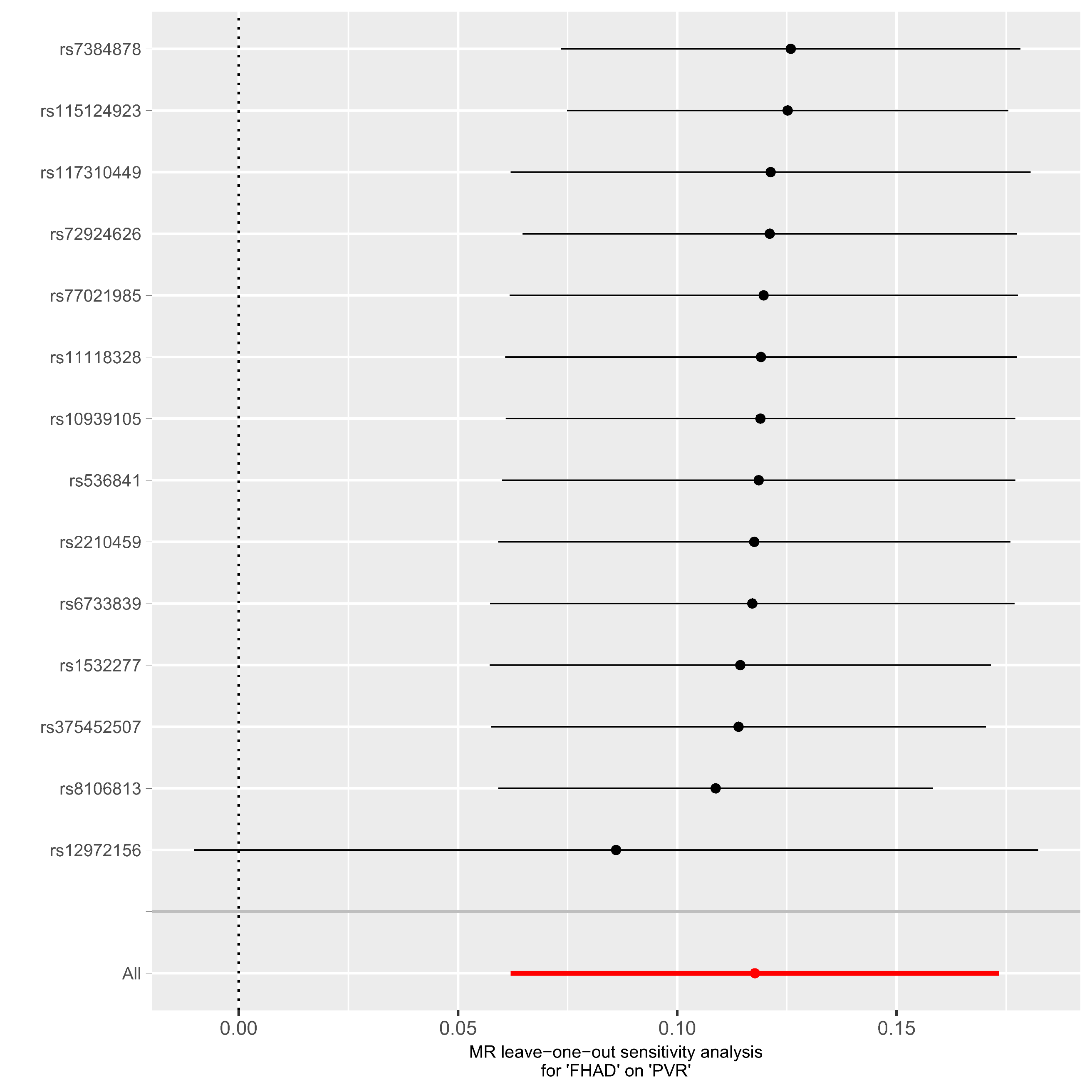
